# Epidemiological inferences from serological responses to cross-reacting pathogens

**DOI:** 10.1101/2024.08.12.24311852

**Authors:** Megan O’Driscoll, Nathanaël Hozé, Noémie Lefrancq, Gabriel Ribeiro Dos Santos, Damien Hoinard, Mohammed Ziaur Rahman, Kishor Kumar Paul, Abu Mohd Naser Titu, Mohammad Shafiul Alam, Mohammad Enayet Hossain, Jessica Vanhomwegen, Simon Cauchemez, Emily S Gurley, Henrik Salje

## Abstract

Multiplex immunoassays are facilitating the parallel measurement of antibody responses against multiple antigenically-related pathogens, generating a wealth of high-dimensional data which depict complex antibody-antigen relationships. In this study we develop a generalizable analytical framework to maximise inferences from multi-pathogen serological studies. We fit the model to measurements of IgG antibody binding to 10 arboviral pathogens from a cross-sectional study in northwest Bangladesh with 1,453 participants. We used our framework to jointly infer the prevalence of each pathogen by location and age, as well as the levels of between-pathogen antibody cross-reactivity. We find evidence of endemic transmission of Japanese encephalitis virus as well as recent outbreaks of dengue and chikungunya viruses in this district. Our estimates of antibody cross-reactivity were highly consistent with phylogenetic distances inferred from genetic data. Further, we demonstrated how our framework can be used to identify the presence of circulating cross-reactive pathogens that were not directly tested for, representing a potential opportunity for the detection of novel emerging pathogens. The presented analytical framework will be applicable to the growing number of multi-pathogen studies and will help support the integration of serological testing into disease surveillance platforms.

## Introduction

Traditional infectious disease surveillance methods rely on testing of symptomatic individuals. However, the number of confirmed cases is often a poor indicator of the true underlying burden of infection in a population due to varying rates of subclinical infections, non-specific symptoms, and heterogeneity in health seeking behaviour (*1*, *2*). Population serological studies, which test for immune markers such as antibodies that are generated in response to an infection, can provide a direct measure of the underlying burden of infection in a population (*3–6*). These studies can help to fill the gaps left by traditional disease surveillance methods, allowing an increased understanding of population susceptibility, pathogen transmission dynamics and rates of pathogen severity (*7–9*).

Recent advances in high-throughput immunoassay technologies have been facilitating the generation of large volumes of immunological data. In particular, multiplex immunoassays which use colour-coded beads to simultaneously measure multiple analytes from a single sample, offers an efficient approach to maximising the information gained from biological samples. In the context of population serological studies, this is leading to a shift in the design of such studies from focusing on a single antigen of interest to now being able to simultaneously test for the presence of antibodies against multiple antigens (*10–18*). Allowing efficient characterisation of population immune profiles against multiple pathogens or antigens, such approaches are paving the way for integrated serosurveillance efforts, providing important new insights for guiding optimal strategies for the control and prevention of infectious agents (*19–21*).

Antibody cross-reactivity has long posed a challenge to the interpretation of serological data in regions where antigenically related pathogens co-circulate (*22*, *23*). In these contexts it can be difficult to determine if antibodies measured against the pathogen of interest were caused by an infection with that specific pathogen or by a related pathogen with similar structural antigenic regions that the antibody can recognize and bind to. Measurements of antibody responses against multiple antigenically-related pathogens can generate a wealth of high-dimensional data depicting complex antibody-antigen relationships, providing a pathway to characterising these cross-reactive dynamics. For instance, the level of correlation in antibody responses measured against different antigens at the population level can provide an indication of their antigenic similarity. Analytical methods that quantify and account for these cross-reactive relationships can maximise inferences from such high-dimensional datasets, allowing robust characterisation of population immune profiles and underlying infection burdens. In addition, cross-reactivity between pathogens provides the potential opportunity to detect novel emerging pathogens via indirect cross-reactive signals within a particular viral family that are otherwise not possible to directly test for.

Arboviruses, including flaviviruses and alphaviruses, are key examples of diverse viral families that represent significant ongoing threats to human health. Members of these viral families continue to emerge into the human population, while those that are already established in human transmission cycles continue their geographic expansion, causing significant burdens to healthcare systems (*24*). Monitoring of the infection prevalence and understanding the antigenic landscape of such viral pathogens is of key importance to global health. In this study, we developed an analytical framework for the analysis of multi-pathogen serological studies to jointly infer the prevalence of infection for each pathogen and the levels of between-pathogen antibody cross-reactivity. We demonstrate the utility of this framework through application to a cross-sectional serological study of arboviruses in the Chapai Nawabganj district in northwest Bangladesh. While cases of Japanese encephalitis virus are regularly reported in this area and a chikungunya virus outbreak was documented in recent years, the underlying infection prevalence of these and other arboviruses in this district is not well understood (*25*, *26*).

### Determining pathogen presence

We analysed serological samples from 1,453 individuals participating in a cross-sectional study conducted in 2014 in Chapai Nawabganj district, located in northwest Bangladesh (Figure 1A). Individuals were enrolled from 39 communities across all 5 sub-districts with an age range of 2-90 years (SI Figure 1). Concentrations of binding IgG antibodies were measured against antigens of 10 pathogens using a Luminex platform: dengue virus serotypes 1, 2, 3 and 4, (DENV1-4), Japanese Encephalitis (JEV), West Nile (WNV), Yellow fever (YFV), Zika (ZIKV), tick-borne encephalitis (TBEV) and chikungunya (CHIKV) viruses. The domain III of the envelope protein (EIII) was used as the target antigen for each flavivirus, while the E2 protein was used for CHIKV. In addition, samples were tested using a commercial DENV enzyme linked immunosorbent assay (ELISA), that used the whole E protein. Values of median fluorescence intensity (MFI) against each antigen from the multiplex immunoassay were divided by individual-level responses to a background control (SNAP tag), to obtain a relative fluorescence intensity (RFI) measurement. Measured RFIs by antigen and upazila (sub-district) of residence are shown in Figure 1B, with respective population distributions of antibody concentrations. Simple Pearson correlation coefficients calculated for RFI titers against each antigen pair revealed high correlations in titers against JEV and WNV (r=0.85), between DENV1 and DENV3 (r=0.80) and between TBEV and YFV (r=0.74), shown in Figure 1C and SI Figure 2. In contrast, we observe low correlation between RFIs measured against CHIKV and all flavivirus antigens, with correlation coefficients ranging from −0.20 to 0.11.

**Figure 1.**
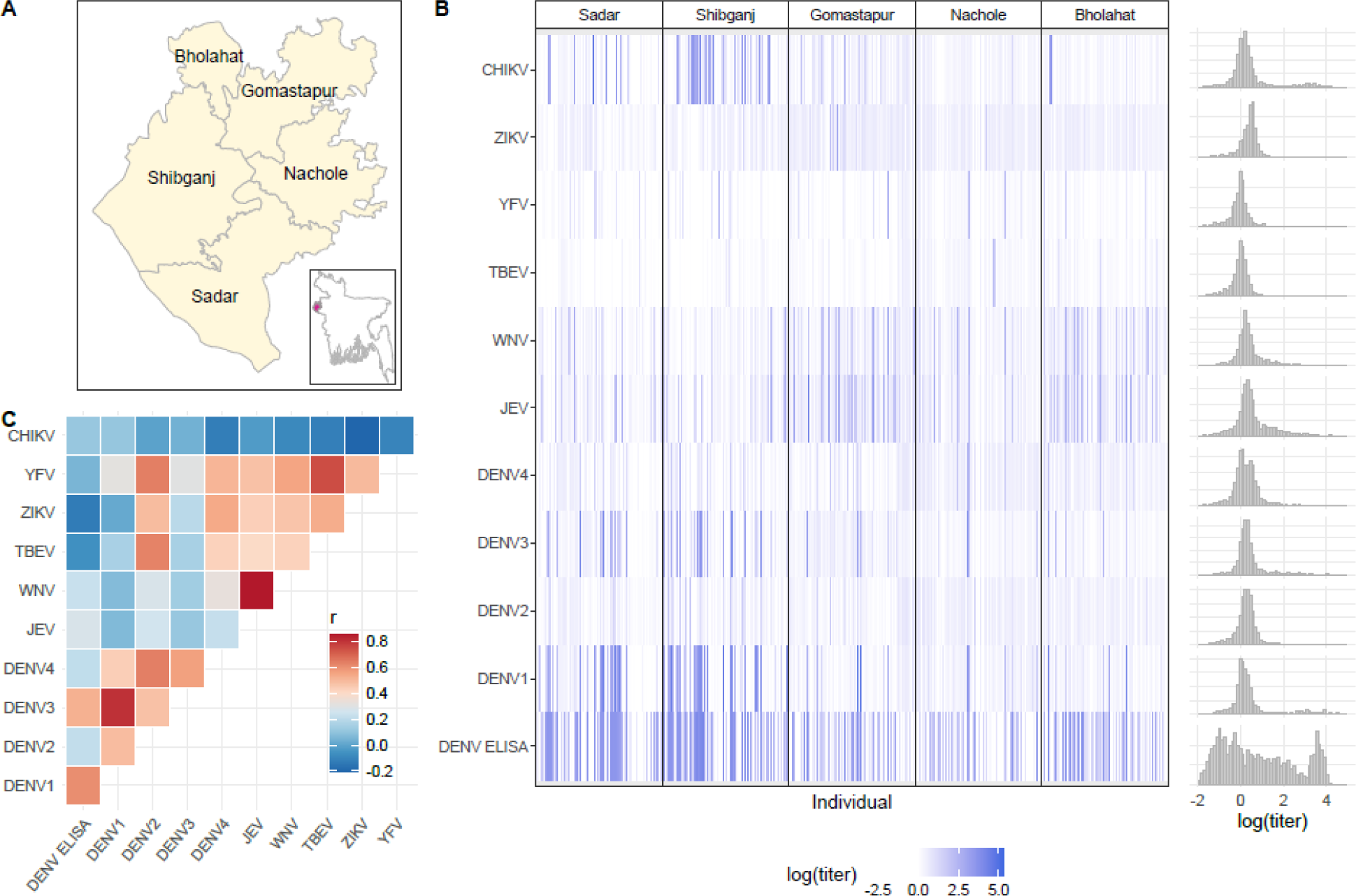
Multi-pathogen serological study data. Panel A shows a map of Chapai Nawabganj district and its 5 sub-districts. The inset plot shows the location of Chapai Nawabganj district within Bangladesh, indicated in red. Panel B shows a heatmap of the log relative fluorescent intensity (RFI) IgG antibody titer values for multiplex antigens as well as log titers for the DENV ELISA assay. Column panels show the antibody binding concentrations by each of the 5 sub-districts. Grey bars on the right show the population distribution of log titer values for each antigen. Panel C shows a heatmap of Pearson r correlation coefficients of population log antibody titers for each multiplex antigen pair.

**Figure 2.**
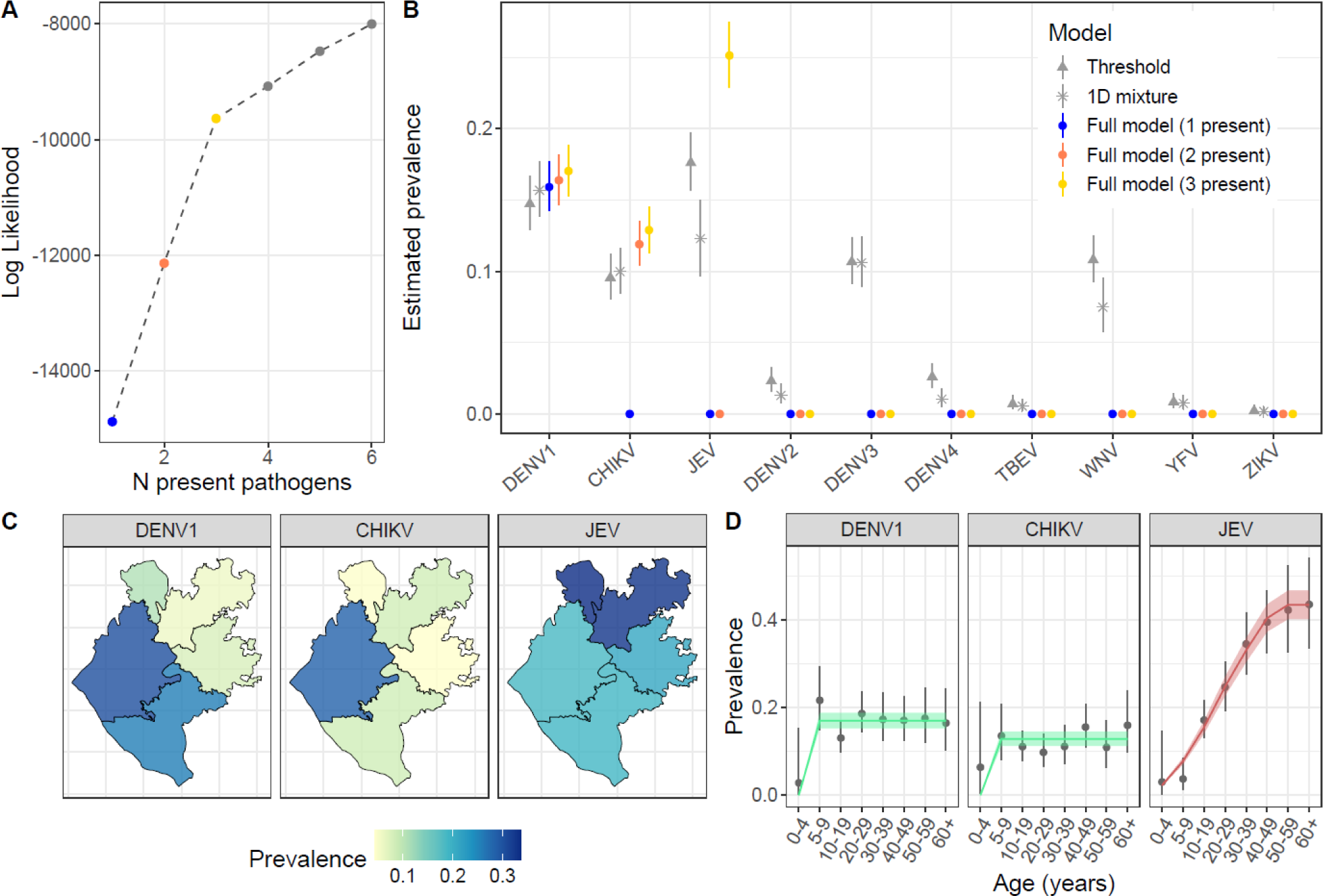
Infection prevalence estimates by space and age. Panel A shows the increase in log-likelihood estimates from the full model as additional pathogens are assumed to be present. Panel B shows a comparison of infection prevalence estimates across models. Points and lines show the median and 95% credible interval estimates of infection prevalence by pathogen. The grey points and lines show estimates from a simple mixture model (stars) and threshold method (triangles) fit independently to antibody data for each pathogen. Coloured points and lines show estimates from the full model framework, progressing from including 1 present pathogen (blue) to 3 present pathogens (yellow). Panel C shows a map of median prevalence estimates by sub-district for each present pathogen from the final full model. Panel D shows the prevalence estimates by age for each present pathogen. Points and lines show the median and 95% credible interval estimates of prevalence from the final multivariate mixture model. The green lines and ribbons show the median and 95% credible interval estimates of population prevalence for DENV1 and CHIKV. The red line and shaded ribbon shows the median and 95% credible interval estimates from a catalytic model for JEV assuming endemic transmission since its estimated introduction time to the region.

We developed a semi-mechanistic multivariate mixture model to jointly infer pathogen-specific infection prevalence and levels of between-pathogen antibody cross-reactivity. In simulation studies we found good model performance on simulated data across a wide range of scenarios (SI). Applying our framework to the arbovirus serological data, we conducted a stepwise variable selection process to determine which pathogens were present, i.e. have transmitted within the study population, and best explain the population antibody titer distributions against all antigens. Fitting to antibody data from all 11 antigens (multiplex immunoassay and ELISA) in the same framework, we first assumed only a single pathogen to be present such that antibody titers against the remaining pathogens can only be explained by negative infection statuses with potential cross-reactivity from the present pathogen. As classic information criterion metrics such as AIC or DIC do not perform well in cases of mixture models of varying components, we instead use likelihood increment percentage (LIP) metrics to assess the relative improvements in model fit with increasing model components (SI methods and simulations) (*27*). Allowing each pathogen to be present in turn, we retained the present pathogen that resulted in the highest model log-likelihood before adding a second present pathogen. We repeated this process of adding present pathogens until the log-likelihood increment percentage per component (LIPpc) fell below 1%. We found highest support for the presence of DENV1 in the study population, followed by CHIKV (LIPpc=9.2%) and JEV (LIPpc=5.0%), shown in Figure 2A. Inclusion of a fourth present pathogen resulted in LIPpc values below 1% and we therefore assumed all the remaining pathogens to be absent from the study population. The selected present pathogens from this variable selection process were consistent across the 3 model versions considered - a base model where a single value of prevalence is estimated by pathogen, a location-specific model where prevalence is estimated by sub-district, and a location and age-specific model where prevalence is estimated by sub-district and age group (SI Tables 4-6). We found the location and age-specific model version performed best by log-likelihood (SI Tables 4-6) and we therefore focus on the results of this model in the remainder of the manuscript. In simulation studies we also found that model versions that allow pathogen-specific prevalence to vary within the population (e.g. by location and/or age) are better able to accurately estimate true parameter values compared to those estimating a single population prevalence per pathogen (SI section 3).

### Inferred infection burden

Our final model estimated an overall infection prevalence of 17.0% (95%CrI: 15.3-18.8%) for DENV1, 12.9% (95%CrI: 11.3-14.6%) for CHIKV and 25.1% (95%CrI: 22.9-27.5%) for JEV in the study population (Figure 2B). We found that pathogen-specific infection prevalence estimates did not significantly change as additional present pathogens were added (Figure 2B). To understand how these estimates would compare to those derived from simpler analytical approaches that are traditionally used, we applied classic single-dimension (1D) mixture models and threshold approaches to antibody data independently for each pathogen. We found infection prevalence estimates for DENV1 and CHIKV to be consistent across models, while classic mixture model and threshold estimates of prevalence for DENV2, DENV4, TBEV, YFV and ZIKV are close to zero, roughly aligned with inferences from our model framework. However, classic mixture models estimated a prevalence of 10.6% (95%CrI: 8.9-12.4%) for DENV3 and 7.5% (95%CrI: 5.7-9.5%) for WNV, inconsistent with our model framework which found these antibody responses to be explained by cross-reactivity from DENV1 and JEV (Figure 2B). In a simulation study we further show how bias in prevalence estimates obtained by classic 1D mixture models increases as both cross-reactivity from a related pathogen and prevalence of the related pathogen increase (SI section 3).

Our model framework estimated substantial spatial variability in infection burden across the district. The prevalence of DENV1 ranged from 5.9% (4.0-8.3%) in Gomastapur in the north of the district to 30.3% (95%CrI: 25.8-34.6%) in Shibganj in the south (Figure 2C). CHIKV prevalence was estimated to range from 4.4% (95%CrI: 2.1-7.8%) in Nachole to 28.4% (95%CrI: 24.3-32.7%) in Shibganj, consistent with where an outbreak had previously been detected (*26*). The prevalence of JEV infection ranged from 18.6% (95%CrI: 14.9-23.0%) in Shibganj to 33.7% (95%CrI: 27.4-40.4%) in Bholahat (Figure 2C). Age-specific prevalence of DENV1 and CHIKV appeared constant among those ≥5 years of age (Figure 2D). The lower prevalence estimated among 0-4 year olds for DENV1 and CHIKV suggests the occurrence of an epidemic for each of these viruses sometime within the 5 years prior to the serological study (2009–2014). In contrast, we observed a pattern of increasing JEV prevalence by age in this district, indicative of endemic transmission (Figure 2D). To characterise the transmission intensity of JEV we fit simple catalytic models to age-specific infection prevalence estimates assuming endemic transmission of JEV since its introduction to Bangladesh. Patterns in the age-specific prevalence estimates suggest the most likely JEV introduction date between 1965-1970 (Figure 2D and SI Figure 3), roughly aligned with the first human cases reported in Bangladesh in 1977 (*28*, *29*). We estimated that 1.2% (95%CrI: 1.1-1.3%) of the susceptible population in this region have been infected each year since its introduction, corresponding to approximately 19,179 (95%CrI: 17,347-21,128) annual JEV infections in the Chapai district (Figure 2D). Estimates of transmission intensity of JEV varied from 0.8% (95%CrI: 0.6-1.1%) in Nawabganj Sadar to 1.7% (1.4-2.0%) in Gomastapur (SI Figure 4).

**Figure 3.**
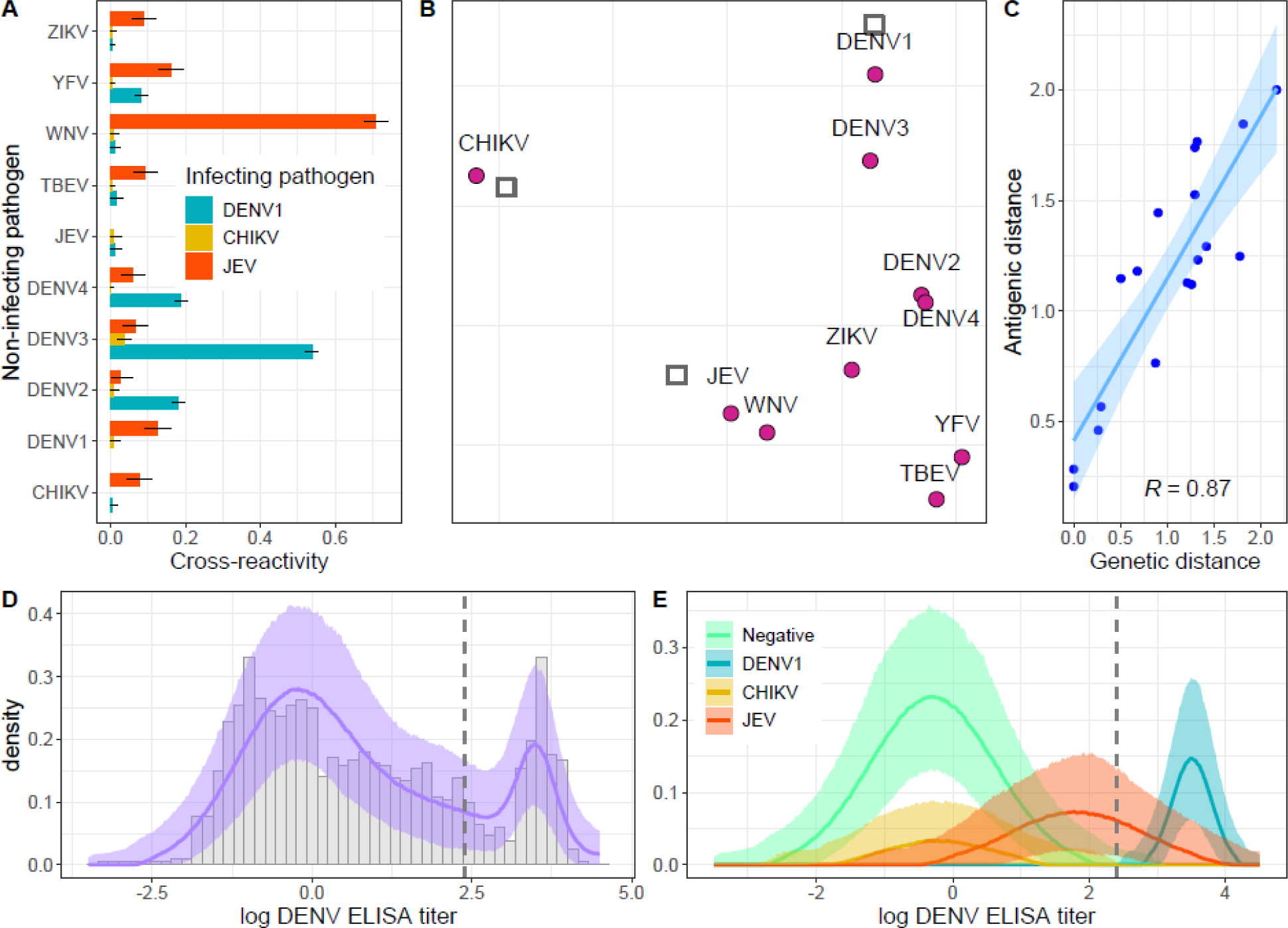
Antigenic relationships. Panel A shows the model estimates of between-pathogen antibody cross-reactivity. Coloured bars show the median model estimates of relative titer increase from each infecting pathogen against each non-infecting pathogen. Black lines show the 95% credible interval estimates. Panel B shows a map of inferred antigenic distances between each sera (grey squares) and antigen (purple circles), obtained by multidimensional scaling of the cross-reactivity estimates to a 2-dimensional space. Panel C shows a comparison of the inferred antigenic distances and genetic distances inferred using phylogenetic methods (blue points), considering only flavivirus pathogen pairs (i.e. excluding CHIKV). The blue line and shaded ribbon show the mean and 95% confidence interval model fit of a linear regression with Pearson correlation coefficient R=0.87. Panel D shows the observed (grey bars) and reconstructed DENV ELISA population titer distribution, where the purple line and shaded ribbon show the model median and 95% credible interval estimates. Panel E shows the reconstructed DENV ELISA titer distributions by infection status where coloured lines and shaded ribbons indicate median and 95% credible interval estimates. The grey dashed line in panels D and E shows the manufacturer recommended threshold for defining DENV seropositivity.

**Figure 4.**
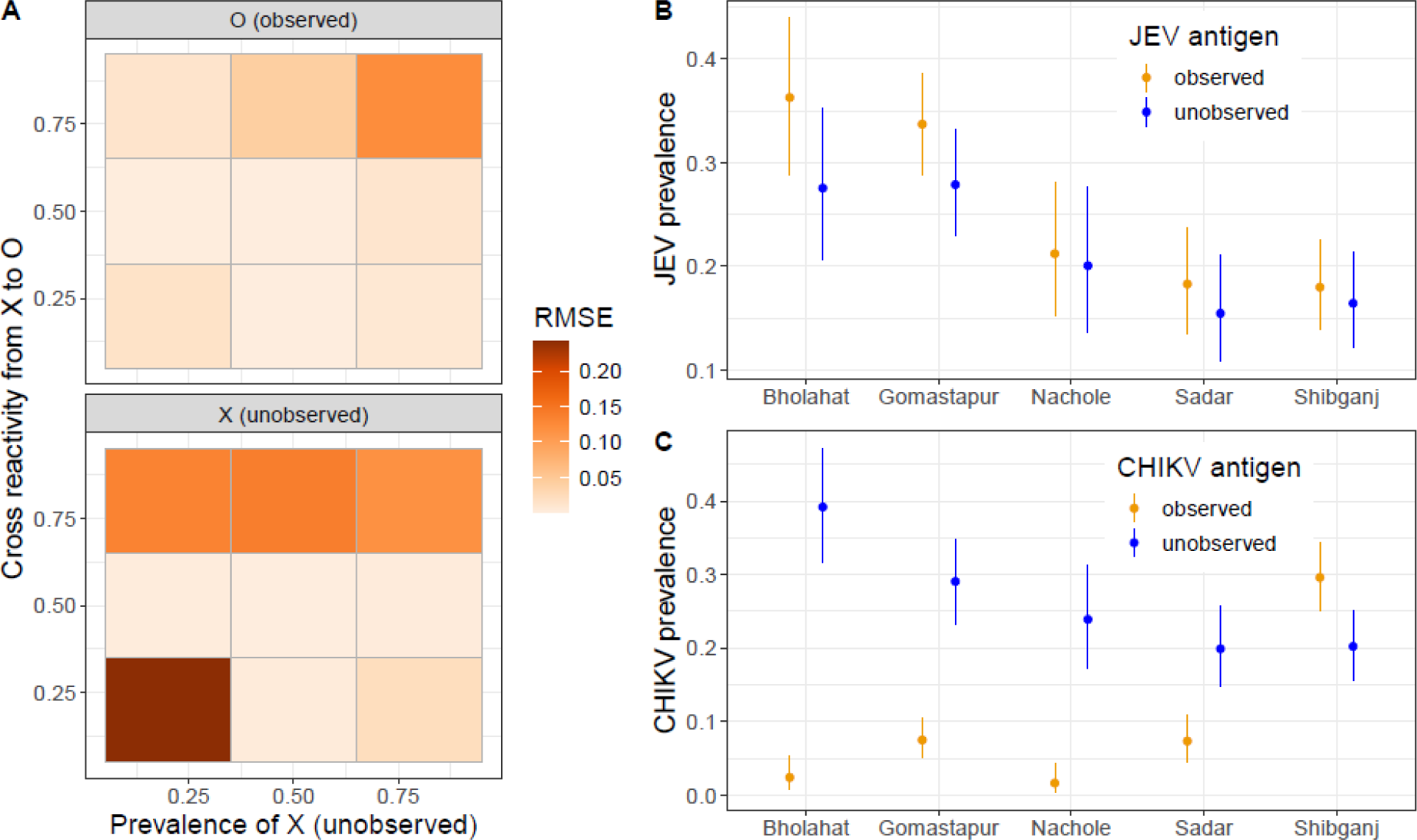
Accounting for unobserved pathogens. Panel A shows a heatmap of the root mean square error (RMSE) of prevalence estimates obtained from simulated data for an observed pathogen, O, and an unobserved pathogen, X, with varying levels of cross reactivity from X to O and varying prevalence of the unobserved pathogen X. Panel B shows a comparison of model estimates of JEV prevalence when antibody titers against the JEV antigen are observed and included in the model (orange) and when they are excluded/unobserved but reconstructed as an unobserved pathogen X. Panel C shows a comparison of model estimates of CHIKV prevalence when CHIKV antibody titers are observed and included in the model (orange) and when they are unobserved/excluded but reconstructed as an unobserved pathogen. Points and lines indicate model median and 95%CrI estimates.

### Between-antigen antibody cross-reactivity

From this best-performing model with three present pathogens we estimated the highest antibody cross-reactivity from JEV to WNV with a relative titer increase of 0.71 (95%CrI: 0.68-0.74) against WNV among JEV positive individuals (Figure 3A). JEV infection was also estimated to induce a 0.16 (0.13-0.19) relative titer increase against YFV and a 0.13 (0.09-0.16) relative titer increase against DENV1. Among DENV1 positive individuals we estimated a 0.54 (95%CrI: 0.52-0.55) relative titer increase against DENV3 as well as a 0.18 (0.16-0.20) and 0.19 (0.17-0.20) relative titer increase against DENV2 and DENV4, respectively (Figure 3A). In contrast, we estimated minimal cross-reactivity from CHIKV infected individuals against the flavivirus antigens, with median estimates ranging from 0.00 to 0.03 (Figure 3A). To summarise the antigenic relationships inferred by the model we applied multidimensional scaling of the estimated antibody responses across infection statuses (Figure 3B). We observed close antigenic clustering of the DENV serotypes, particularly DENV2 and DENV4, though it is worth noting that the positions of these DENV antigens are informed only by the positive sera of a single DENV serotype (DENV1). We also inferred a close antigenic relationship between JEV and WNV, members of the same serocomplex (*23*), as well as between TBEV and YFV (Figure 3B). To understand how our inferred antigenic distances compared to phylogenetic distances within the *Flaviviridae* family, we constructed a maximum-likelihood phylogeny of representative flavivirus E proteins. We found high correlation between our inferred antigenic distances and genetic distances with Pearson r=0.87 (Figure 3C). The shortest genetic and antigenic distances were estimated between JEV and WNV and between DENV1 and DENV3, while the greatest antigenic and genetic distances were inferred between DENV1 and TBEV and between DENV1 and YFV (Figure 3C).

To understand the relationship between each multiplex antigen and the more sensitive DENV ELISA antigen we estimated where the antibody titer distributions of each infection status fall on the DENV ELISA assay. The full reconstructed distribution of DENV ELISA titers is shown in Figure 3D. Of the present pathogens included in our model, we estimated that DENV1 positive individuals have the highest response on the DENV ELISA assay with mean titers of 3.5 (95%CrI: 3.5-3.6), shown in Figure 3E. We also estimated a significant signal from JEV positive individuals to the DENV ELISA assay with mean titers of 1.8 (95%CrI: 1.7-1.9), while CHIKV positive individuals had mean DENV ELISA titers of −0.2 (95%CrI: −0.4-0.0) similar to that of individuals negative to all pathogens (mean DENV ELISA titer −0.3, 95%CrI: −0.4 - −0.2) (Figure 3E). Using the manufacturer recommended cutoff for defining DENV seropositivity results in a prevalence of 22.0% (95%CI: 19.9-24.3%), slightly higher than our estimates of DENV1 prevalence of 17.0% (95%CrI: 15.3-18.8%), inferred by the full model. Results from our model suggest the difference in estimates is attributed to JEV infected individuals that have high responses on the DENV ELISA assay (Figure 3E). We note that this cross-reactivity from endemic JEV leads to a trend of increasing DENV ELISA titers by age (IS Figure 5), which could easily be misinterpreted as DENV endemicity when the DENV ELISA results are taken on their own.

### Detecting unobserved pathogens

A key uncertainty that arises in multi-pathogen serological studies is the potential presence of cross-reactive antibodies induced from unmeasured or unknown pathogens. This raises the potential issue of misattributing antibody responses to related pathogens that have been tested for. In this section we use our analytical method to explore the scenarios in which the presence and prevalence of an unobserved pathogen can be inferred through cross-reactive antibody responses. We conduct simulations of a 3 pathogen system, where 2 pathogens are observed/tested for and 1 is not. We find that estimates of the prevalence of observed pathogens (O) and the unobserved pathogen (X) can both be accurately inferred at intermediate levels of cross reactivity from X to O, shown in Figure 4A, with low root mean square error (RMSE). At lower levels of cross-reactivity from X to O, the prevalence of observed pathogens O can be estimated with good accuracy due to limited interference from X. However, the prevalence of X cannot be well identified when both its true prevalence and its cross reactivity to observed pathogens are both low. At higher levels of cross reactivity from X to O, the prevalence of X cannot be accurately reconstructed, with the prevalence of O also becoming unidentifiable as the prevalence of X increases (Figure 4A).

To assess the generalizability of these findings to real data we refit our final model to the arbovirus antigens, excluding one antigen and attempting to reconstruct the parameters for this unobserved pathogen. We show that our model was able to accurately reconstruct the prevalence of JEV as an unobserved pathogen when measurements from the JEV antigen are excluded from model fitting (Figure 4B). In addition, the model accurately reconstructed estimates of cross reactivity from the unobserved JEV antigen to each observed antigen, including its close antigenic relationship to WNV (IS Figure 6). We note that this model performed better by log-likelihood compared to when WNV is assumed to be the third present pathogen. In contrast to the case of JEV, we show that when the CHIKV antigen is excluded from fitting, the model was not able to accurately reconstruct the parameters of CHIKV. This is consistent with the simulation results that demonstrate the lack of cross reactivity between CHIKV and flavivirus antigens inhibits its identification while the intermediate level of cross reactivity from JEV to the DENV ELISA facilitates the accurate reconstruction of JEV parameters.

## Discussion

The transmission of multiple antigenically related pathogens in a population has long posed a challenge to the interpretation of serological studies of flaviviruses and other antigenically-variable pathogens. In contexts where >1 related, cross-reacting pathogens have been transmitted in a population, it can be difficult to understand if antibodies detected against a specific pathogen were induced through exposure to that or a related pathogen. In this study we applied an analytical framework to multi-pathogen serological data to disentangle the multi-dimensional antigen-antibody relationships. We used this framework to infer estimates of arboviral prevalence in northwest Bangladesh by location and age, accounting for varying levels of between-pathogen antibody cross-reactivity.

A key question arising from multi-pathogen serological studies is determining which pathogens have truly transmitted in the study population. To date in Bangladesh, there has been no evidence of human transmission for YFV or TBEV, while human cases of DENV, CHIKV, JEV, and more recently ZIKV and WNV have all been reported in the country (*25*, *26*, *28*, *30–32*). Among the pathogens included in the study we found evidence of past DENV, CHIKV and JEV transmission in the district of Chapai Nawabganj, while no strong evidence for the presence of the remaining pathogens was found. We show that simpler methods that do not account for antibody cross-reactivity would have inferred the presence of WNV, while in our model WNV antibodies are found to be explained by high cross-reactivity from JEV infections. This is consistent with local epidemiological data, with confirmed JEV hospitalised cases regularly reported in this district (*25*). In contrast, the first diagnosed human case of WNV in Bangladesh was reported in 2019 in Dhaka, 5 years after the completion of the serological study (*32*). We infer the highest prevalence of DENV and CHIKV in the more urbanised district of Shibganj, where a past epidemic of CHIKV had been reported and consistent with the *Aedes* vector’s ability to thrive in urban environments (*26*). For ZIKV, the first detected human case in Bangladesh was in August 2014 with serological evidence of ZIKV infections in Dhaka since 2013 (*30*, *33*). However, we found no evidence of ZIKV transmission in the district of Chapai Nawabganj as of 2014.

We find our model estimates of infection prevalence to be consistent with or without including the DENV ELISA assay in model fitting (IS Figure 7). Estimates of JEV prevalence however, were slightly higher when the DENV ELISA assay was included in model fitting, consistent with the lower sensitivity reported for the multiplex JEV EDIII antigen used in this study (*34*) and the high sensitivity of a whole E protein DENV ELISA assays. Our analysis explored how this combination of sensitive and specific antigens can be leveraged as a means of detecting the presence and prevalence of unknown or unmeasured pathogens. We demonstrated how signals from related cross reacting pathogens can make the detection of unobserved pathogens possible under conditions of intermediate cross reactivity profiles. Future work to define robust criteria or strength of evidence for the detection of unobserved pathogens through serological surveillance will be important for public health applications. The major emerging pathogens of recent years have all belonged to families of known human pathogens, with coronaviruses, flaviviruses and influenza viruses being the predominant examples. Our findings highlight how novel emerging pathogens could be detected through routine serological testing for a range of pathogen families, adding support to the potential benefits of a global immunological observatory and routine serological surveillance (*35*, *36*).

There are a number of limitations that should be considered when interpreting the results of our analyses. Antibody kinetics are not accounted for in our model framework which relies on cross-sectional serological data representing a snapshot of population antibody titers at a single point in time. As the timing and sequence of infections cannot be accounted for, our model estimates will therefore reflect population averages across a range of time-since-infection scenarios represented in the study population. If antibodies wane significantly following infection then the prevalence of infection may be underestimated. However, in the case of the arboviruses measured in this study, infection is thought to induce long-term homotypic IgG responses (*37*, *38*). In addition, if an infection induces higher antibody responses against a related pathogen than the infecting pathogen then the infection statuses may be misclassified. We assume that cross reactive antibodies from infecting pathogens to any non-infecting pathogen combine additively which may not hold for individuals infected with an increasing number of related pathogens. Our model represents an unsupervised approach to classifying population infection statuses which has the advantage of not relying on validated sample sets that can be time- and resource-intensive to obtain. While our extensive simulation study demonstrated the theoretical validity of our analytical framework (SI), future analysis from serological cohort studies where infections are confirmed by PCR or other methods will allow an improved understanding of model performance.

Despite these limitations, a comparison of antigenic distances between assay antigens and monotypic sera inferred by our model were found to be highly consistent with genetic distances between antigens independently inferred from phylogenetic methods. In addition, our inferences of arbovirus infection burden are consistent with known epidemiological indicators of each present pathogen. Previous studies have analysed serological data considering two related pathogens using additional assumptions of pathogen transmission dynamics (*39*, *40*). The analytical framework presented here is agnostic to any specific pathogen transmission dynamics allowing its scalability to many pathogens with potentially different transmission dynamics. This framework has shown to provide robust estimates of infection prevalence in systems of several related pathogens while simultaneously accounting for varying levels of between-pathogen cross-reactivity.

In this study we demonstrate the utility of this novel analytical approach for making robust epidemiological inferences of public health relevance. This framework was able to disentangle high-dimensional antibody-antigen relationships of multiple related pathogens, providing new insights into the burden and transmission dynamics as well as the immune landscape of arboviral diseases.

## Data Availability

Simulated data and de-identified data are available in the associated GitHub repository

https://github.com/meganodris/MultiSero

## Disentangling the dynamics of cross-reacting pathogens in serological studies: Supplementary Material

### 1 Methodology

#### 1.1 Serological study

We analyse data from a cross-sectional seroprevalence study conducted in 2014 in the Chapai Nawabganj zila (district) of Bangladesh, located in the Rajshahi division. A total of 1,453 individuals were included in the study from 39 communities across the 5 upazilas (sub-districts) of Chapai Nawabganj. Participant recruitment in each rural community was initiated by identifying the household where the most recent wedding took place and beginning recruitment in the house of their closest neighbour. In each urban community, recruitment was initiated by finding the nearest community center and identifying the closest neighbouring household for participant recruitment. All residents of selected houses, regardless of age, were eligible for enrolment in the study. After the first enrolled household, the following 5 closest neighbours were skipped and the 6th closest household was approached for enrollment. This process was repeated until individuals from at least 15 different households had been enrolled from that community. Individuals that were unable to give consent due to disability or who had an acute medical condition where blood collection is contraindicated were excluded from study participation. A single 5ml venous blood sample was collected from each enrolled participant. The age distribution of the study participants, compared to the population age distribution for each of the 5 upazilas is shown in Figure 1. Data on the population age distributions were obtained from the 2011 Bangladesh census [1].

**Figure 1.**
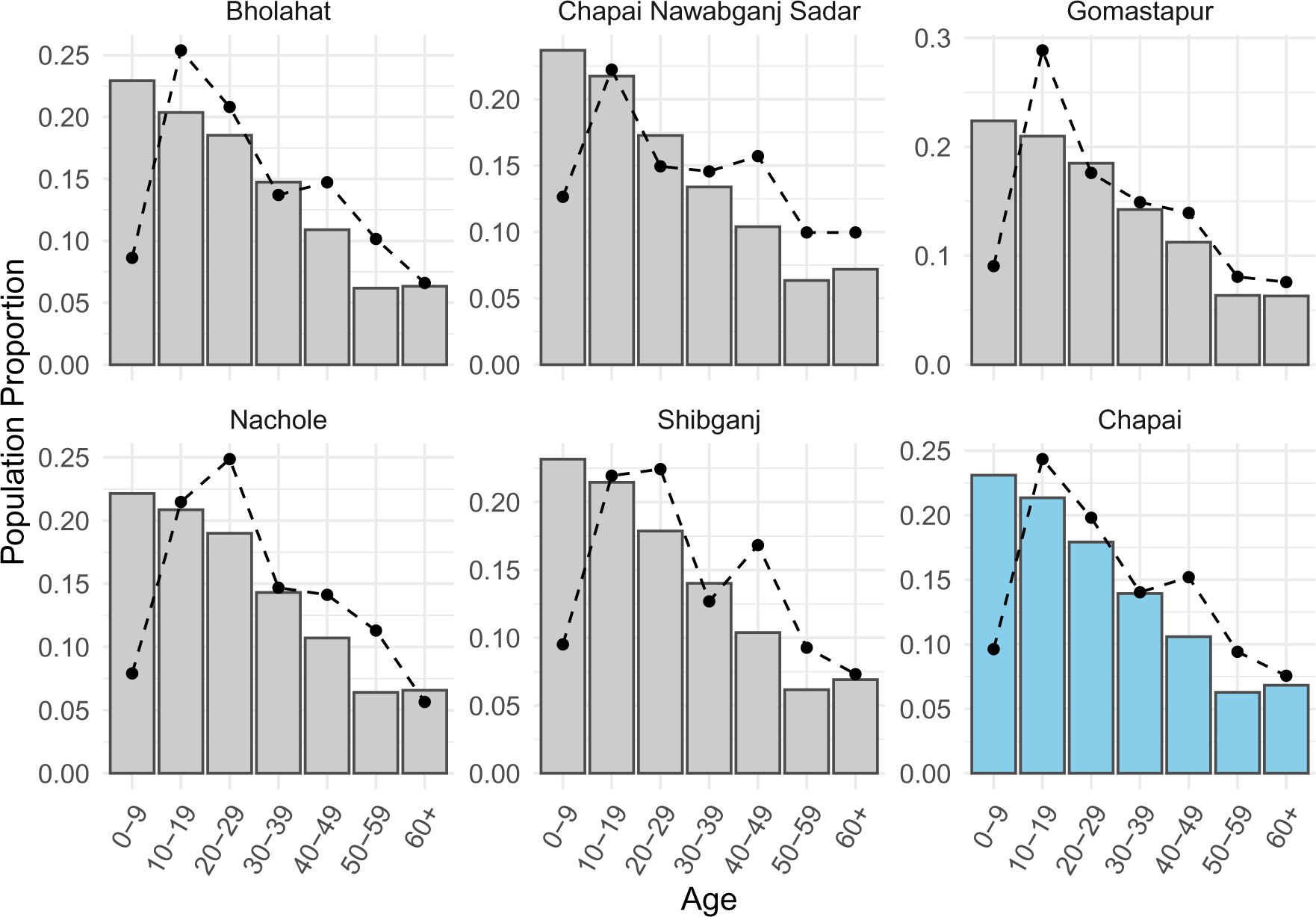
Age distribution of study participants. The grey bars in each panel show the population age distribution for each upazila and the blue bars show the population age distribution for the wider zila of Chapai Nawabganj (Chapai). Black points and dashed lines indicate the proportion of study participants in each age group by upazila and across the total study population representing Chapai.

#### 1.2 Multiplex immunoassay

Serum samples were tested for the presence of IgG antibodies against 10 antigens using an in-house microsphere-based multiplex immunoassay (MIA). Recombinant EDIII antigens were used for all flavivirus pathogens (DENV1, DENV2, DENV3, DENV4, JEV, WNV, TBEV, YFV, ZIKV) to improve assay sensitivity and specificity, as previously described [2], while a recombinant E2 glycoprotein was used for CHIKV. Magnetic beads were coupled to each specific antigen and the assay was conducted as previously described including individual-level background controls using a recombinant human protein SNAP-tag (*O*^6^-methylguanine DNA methyltransferase) [2]. Fluorescence intensity and bead colour coding were measured using a Luminex 200 system (Bio-Rad Laboratories). For each sample a relative fluorescence intensity (RFI) value was calculated by dividing the median fluorescence intensity (MFI) by the fluorescence intensity of the individual-level background control. All analyses of MIA data were conducted using log RFI values. In addition to the 10 antigens measured with the MIA, a Panbio DENV ELISA assay that utilises the whole DENV E protein was also used to measure binding DENV IgG antibody concentrations.

#### 1.3 Multivariate mixture model framework

We develop and apply a semi-mechanistic multivariate Gaussian mixture model to jointly infer the prevalence of infection of multiple pathogens and the levels of between-pathogen antibody cross-reactivity. We assume long-term persistence of IgG antibodies following infection and therefore define positivity as the presence or absence of serological evidence of a past infection. We consider a system of *n* = *P* pathogens, where the true infection status of each individual is either negative, {0}, or positive, {1}, for each pathogen. For a system of *P* pathogens with two possible outcomes there are 2*^P^* possible infection status combinations. Table 1.3 shows the example of all possible infection status combinations for a system of 2 and 3 pathogens, A, B and C, with each row corresponding to a unique infection status combination. For each possible infection status combination, *c*, we define a *P−*dimensional Gaussian component, with vector of means, *µ_c_*, and covariance matrix Σ*_c_*. The proportion of the study population with infection status combination *c*, *θ_c_*, is defined as the conditional probability of having infection status combination, *c*, given the pathogen-specific population prevalence, *π_p_*. For example, for infection status combination *c* = {0, 1, 0} where individuals have been infected by pathogen B only, the proportion of the population with this infection status is calculated as *θ_c_* = (1 *−π_A_*)(*π_B_*)(1 *−π_C_*).

**Table 1.**
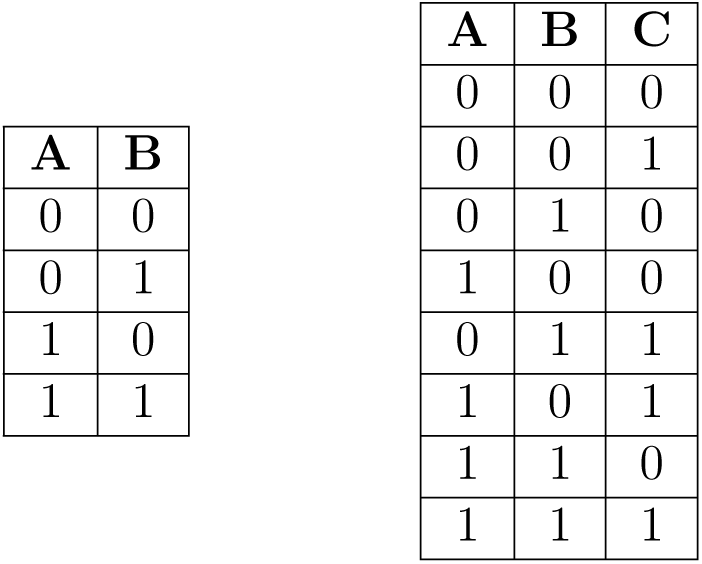
Infection status combinations. All possible infection status combinations for a 2-pathogen and 3-pathogen system, respectively. Each row represents a unique infection status combination for pathogens A, B and C, where 0 represents a negative infection status and 1 represents a positive infection status for each pathogen.

##### 1.3.1 Two pathogen system

We first consider a system of 2 related pathogens, A and B. For each of the 4 possible infection status combinations (Table 1.3), we define a bivariate Gaussian component to characterize the antibody titers of this status. For individuals that are negative to both A and B, *{A*_0_*, B*_0_*}*, the Gaussian component will be defined with means, 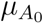 and 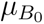, standard deviations, 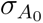 and 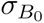. We allow for correlation in these negative antibody titers, with correlation coefficient *ρ*_0_. The covariance matrix, Σ, for this Gaussian component is then defined as shown in equation 1, where the covariance of A and B, 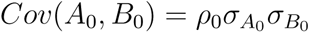.

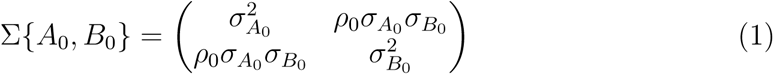

For individuals who have been infected with pathogen B only, {*A*_0_*, B*_1_}, we define the mean rise in antibody titers against pathogen B as 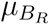, such that the mean antibody titers against pathogen B will be 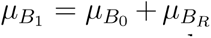, with standard deviation 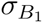. We assume this bivariate Gaussian component to be a linear combination of negative antibody titers against pathogen A and positive antibody titers against pathogen B. In this way the mean antibody titers against pathogen A are calculated as 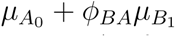. Here, *ϕ_BA_* is the relative increase in antibody titers raised against pathogen A after infection with pathogen B, relative to the antibody titers binding to pathogen B. The covariance in antibody titers for this Gaussian component is calculated as shown in equations 2-3 using the bilinearity property of covariance. Here we assume the covariance in these antibody titers in the absence of cross-reactivity to be zero, *Cov*(*A*_0_*, B*_1_) = 0. Under this assumption, the standard deviation for antibody titers against pathogen A can be calculated as 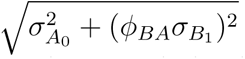. The covariance matrix for this bivariate Gaussian component can subsequently be defined as shown in equation 4.

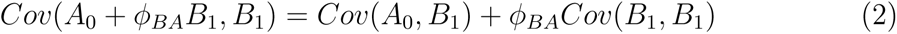

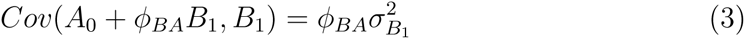

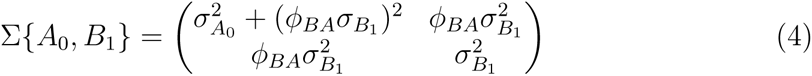

For individuals infected with A only, {*A*_1_*, B*_0_}, the bivariate Gaussian component is defined with the same approach as before. The Gaussian means are defined as 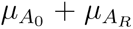 and 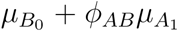, respectively for antibody titers against pathogens A and B, and the standard deviations as 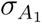 and 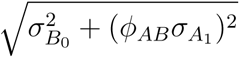. With the same approach as above (equations 2-3), the covariance matrix of this Gaussian component is defined as shown in equation 5. We assume that cross-reactive antibodies cannot decrease antibody titers, i.e. *ϕ ≥* 0. In addition, we allow for asymmetric cross-reactive antibody responses such that *ϕ_BA_* is independent of *ϕ_AB_*.

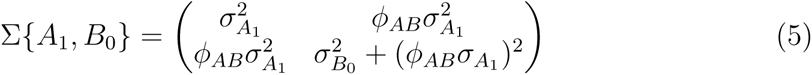

Finally, for infection status combination {*A*_1_*, B*_1_} where individuals have been infected with both pathogens A and B, we define the means of the Gaussian component as 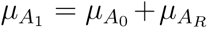 and 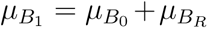. Without knowing the sequence or timing of infections, we assume the means and standard deviations of positive antibody titers to be constant across infection statuses. Therefore, the standard deviations of this Gaussian component are simply defined as 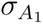 and 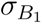 with correlation coefficient, *ρ*_1_. The covariance matrix for this Gaussian component is then defined as shown in equation 6.

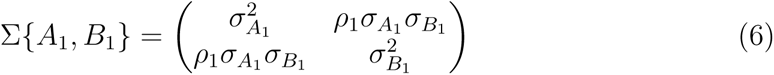

##### 1.3.2 Beyond two pathogens

For systems with increasing numbers of pathogens, *n* = *P*, the number of possible infection status combinations, and therefore the number of *P−*dimensional Gaussian components required to describe the data, increases as 2*^P^*. To reduce the number of new parameters required when scaling the system to higher dimensions, we assume that correlation in antibody titers when individuals are negative to all pathogens, *ρ*_0_, is constant across pathogen pairs. For instance, the covariance matrix for infection status {*A*_0_*, B*_0_*, C*_0_}, in a 3 pathogen system will be defined as shown in equation 7.

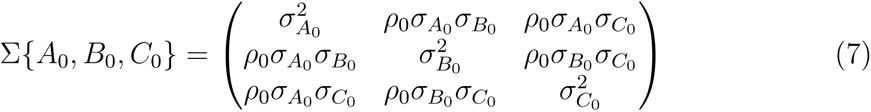

For infection statuses where individuals have been infected with 1 pathogen, the covariance in antibody titers against the infecting pathogen and non-infecting pathogens is calculated with the same approach as before (equations 2 - 3). The co-variance in antibody titers for pairs of non-infecting pathogens, now in the presence of cross-reactive antibodies from a related pathogen, is calculated as shown in equations 8 - 10 taking the example of infection status {*A*_0_*, B*_0_*, C*_1_}, where individuals are positive to pathogen C only. As before, we assume antibody titers against infecting and non-infecting pathogens to be independent in the absence of cross-reactivity such that *Cov*(*A*_0_*, C*_1_) = 0 and *Cov*(*B*_0_*, C*_1_) = 0. The covariance matrix for this infection status is then defined as shown in equation 11.

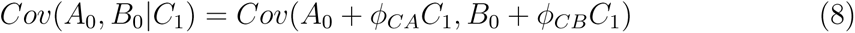

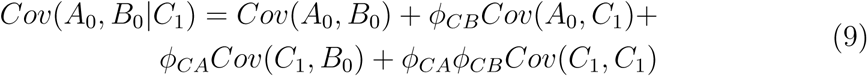

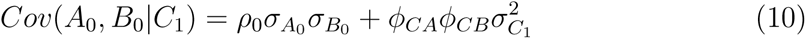

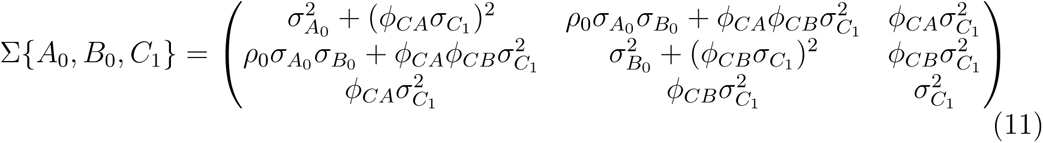

In the case of infection status combinations where individuals have been infected with *>* 1 pathogens, we assume the means and standard deviations of cross-reactive antibody titers against non-infecting pathogens to increase additively. For instance, for an infection status {*A*_0_*, B*_1_*, C*_1_}, the mean of the Gaussian component characterising titers against pathogen A will be calculated as 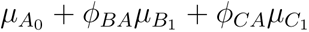. Similarly, the standard deviation for antibody titers against pathogen A is calculated as 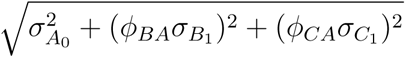. The covariance between negative and positive titers is now calculated as shown in equation 12, additionally accounting for the correlation in antibody titers for the positive pathogens, *ρ*_1_. We assume this correlation in antibody titers between pairs of positive pathogens to be constant across all pathogen pairs and infection status combinations. The covariance matrix for infection status {*A*_0_*, B*_1_*, C*_1_} is shown in equation 13.

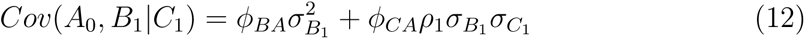

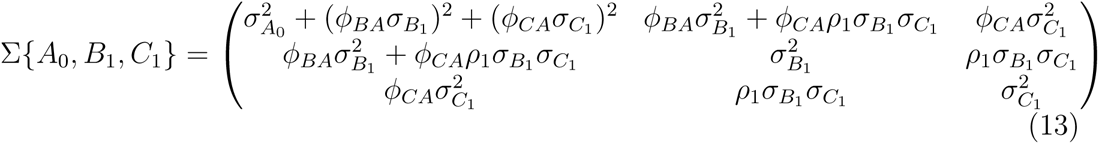

More generally, in the case of covariance in antibody titers against pairs of non-infecting pathogens, in the presence of *>* 1 infecting pathogen, the covariance is calculated as shown in equation 14. In the case of covariance in antibody titers for an infecting and non-infecting pathogen, in the presence of *>* 1 infecting pathogen, the covariance is calculated as shown in equation 15.

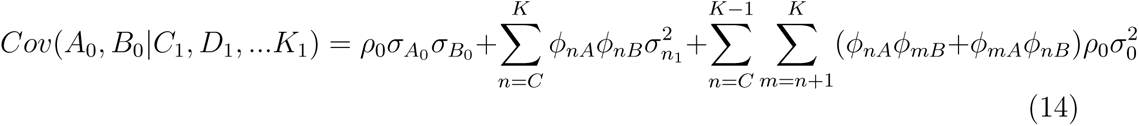

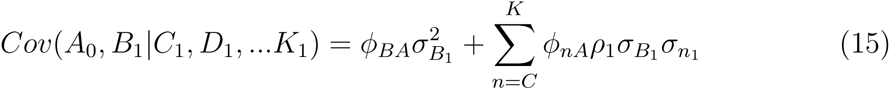

##### 1.3.3 Present vs absent pathogens

To understand which pathogens are likely to have transmitted in the population we conduct a step-wise variable selection process, comparing the performance of models that assume a pathogen to be present vs absent. In the case of pathogens that are assumed to be absent from the study population, the model can be simplified by reducing the number of possible infection status combinations and therefore the number of Gaussian components needed to describe the data. For instance, considering a system of 3 pathogens, A, B and C, there are 8 possible infection status combinations if all 3 pathogens have transmitted in the population. In contrast, if pathogen C can be assumed to be absent from the study population (i.e. has never transmitted in the population) then the number of infection status combinations is halved, as shown in Table 1.3.3. In this way the antibody titers against the absent pathogen are still included in model fitting and are characterised only by the negative and negative with cross-reactivity Gaussian components/infection statuses. The number of parameters is also reduced, with *π* and *µ*_1_ parameters no longer estimated for each absent pathogen. Cross-reactivity parameters *ϕ* from the absent pathogens to present pathogens will also not be estimated but cross-reactivity from the present pathogens to the absent ones are estimated.

**Table 2.**
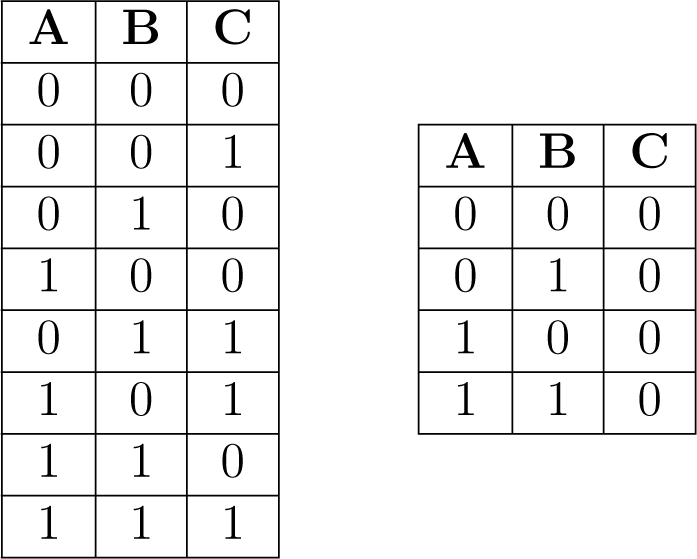
Infection status combinations & absent pathogens. All possible infection status combinations for a 3-pathogen system where all pathogens are present and where only pathogens A and B are present, respectively.

##### 1.3.4 Unobserved pathogens

We explore the ability of the model framework to accurately reconstruct the infection prevalence of some unmeasured/unobserved pathogen, “pathogen X”, that has not been tested for by the serological assay. Additional Gaussian components are fit to characterize the antibody titer distributions of pathogen X, with 2*^P^* ^+1^ possible infection status combinations. Covariance matrices for each infection status combination will be of size *P ∗ P*, where *P* is the number of observed pathogens/antigens. The prevalence of the unmeasured pathogen is estimated as well and the cross-reactivity from the unobserved to the measured pathogens. As the homologous binding to pathogen X is unknown, we assume some fixed value of mean titers against pathogen X, from which the relative titer increase against other pathogens can be inferred. We assume the standard deviation of these positive titers against pathogen X to be equal to the standard deviation of positive responses against the measured pathogens.

#### 1.4 Model fitting

We applied the model framework to the arbovirus serological data from Chapai Nawabganj. We considered the data as being representative of 10 pathogens that were tested using the MIA (CHIKV, DENV1, DENV2, DENV3, DENV4, JEV, TBEV, WNV, YFV, ZIKV) and an additional antigen (DENV ELISA assay) giving a total of 11 antigens. We assumed the standard deviation of negative antibody responses, *σ*_0_, and positive antibody responses, *σ*_1_, are each constant across the multiplex immunoassay antigens. We also assumed that correlation in antibody responses against pairs of infecting pathogens, *ρ*_0_, to be 0. We fit the multivariate mixture model framework jointly to data from all 11 antigens and conducted a step-wise variable selection process to identify which of the 10 pathogens are likely to be present in the study population. We considered a “base” model version that estimates a single infection prevalence per pathogen, as well as location-specific and location- and age-specific versions. In the location-specific versions, pathogen-specific prevalence is allowed to vary by each of the 5 sub-districts of Chapai Nawabganj. For the location- and age-specific version we further allowed pathogen prevalence to vary by age group. We assumed all other parameters to be constant across the study population, allowing only pathogen-specific infection prevalence to vary within the population. All code used for model fitting is available at https://github.com/meganodris/MultiSero.

##### 1.4.1 Variable selection process

We started by assuming only 1 of the 10 pathogens to be present in the study population, such that antibody titers for the remaining 9 pathogens are forced to be explained by negative infection statuses and cross-reactive antibodies from the single present pathogen. We fit this model assuming each of the 10 pathogens to be present in turn and identified the model with the highest log-likelihood from this step. The present pathogen from the best performing model was then retained in the next step where a second present pathogen is added. We repeated this iterative process, at each step identifying the additional present pathogen that improved the likelihood the most. We continued the process until the likelihood increment percentage per component (LIPpc) fell below a threshold of 1%. Due to the limitations of traditional information criterion approaches for the comparison of finite mixture models of varying complexity, we focused on the likelihood increment percentage (LIP) metric [4, 6]. We used the LIPpc to assess the improvement in model fit across models with an increasing number of present pathogens and therefore increasing numbers of Gaussian components used to explain the data. Here the increase in log-likelihood for model *j*, relative to the log-likelihood of simpler model *j−* 1 is considered, accounting for the additional Gaussian components, *g*, fitted by model *j*, *g_j_*, shown in equation 16.

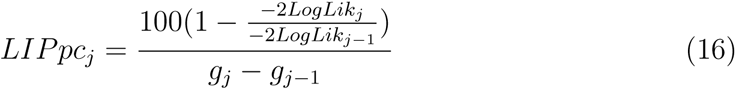

##### 1.4.2 DENV ELISA

We included the DENV ELISA as an additional antigen/dimension in the full model framework. Due to the inclusion of DENV antigens in the MIA, we did not consider the DENV ELISA as an independent pathogen that can be present. We instead allowed the DENV ELISA antibody titers to be explained by the same Gaussian components describing the MIA antibody responses. We assumed no covariance between antibody titers measured on the DENV ELISA and on the MIA. We estimated the mean, *µ_E_*, and standard deviation, *σ_E_*, of DENV ELISA titers for infection status combinations that are negative to all pathogens and those positive to a single pathogen. For infection statuses that are positive to *>* 1 pathogen we assumed the mean DENV ELISA titers to be equal to the maximum *µ_E_* of the Gaussian components describing the respective monotypic infection statuses. The standard deviation of DENV ELISA titers for an infection status positive to *>* 1 pathogen is calculated as the joint standard deviation of the respective monotypic infection statuses. For example, the standard deviation of DENV ELISA titers for an infection status positive to pathogens A and B, 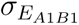, was calculated as 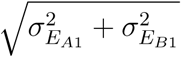.

##### 1.4.3 Reconstructing unobserved pathogens

To test the ability of our model to reconstruct the infection prevalence of an unobserved pathogen, we exclude antibody titer data from a single pathogen from model fitting. In particular, of the pathogens that were determined to be present in the study population from the variable selection process, we excluded the multiplex antigen of each in turn. We included the DENV ELISA assay in model fitting and inferred infection prevalence of each observed and unobserved pathogen by sub-district. We assumed a fixed value of 2 for mean positive titer responses against pathogen X, for inference of the cross reactivity responses from pathogen X to observed pathogens.

##### 1.4.4 Likelihood & Priors

We assumed that the antibody titers for each infection status combination follow a multivariate Gaussian distribution, with the probability of observing antibody titers *x* for individual *i*, given infection status combination *c* shown in equation 17. Here, *x* is a *n* = *P* vector of antibody titer measurements to each pathogen. The full model log-likelihood is calculated as the sum of the log probabilities across individuals, *i*, and across all possible infection status combinations, *c*, weighted by the proportion of the study population with infection status combination *c*, *θ_c_*, shown in equation 18. The log likelihood of the location and age-specific model is calculated similarly to before, shown in equation 19, but now with location- and age-specific *θ_c,l,a_* derived from the location- and age-specific prevalence estimates *π_p,l,a_* as previously described.

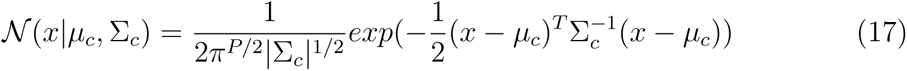

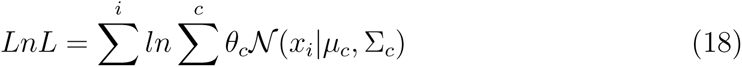

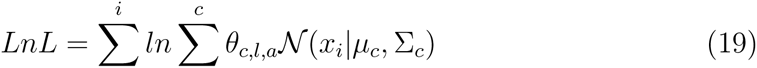

Parameter priors are shown in Table 3, which were assumed to be constant across pathogens. We fit the model in a Bayesian framework with Hamiltonian Monte Carlo No-U-Turn sampling using cmdStanR [9]. Each model was fit with 3 chains for 3,000 iterations in addition to 3,000 warm-up samples. Model convergence was assessed by visual inspection of chain mixing and by R-hat convergence diagnostic across all parameters.

**Table 3.**
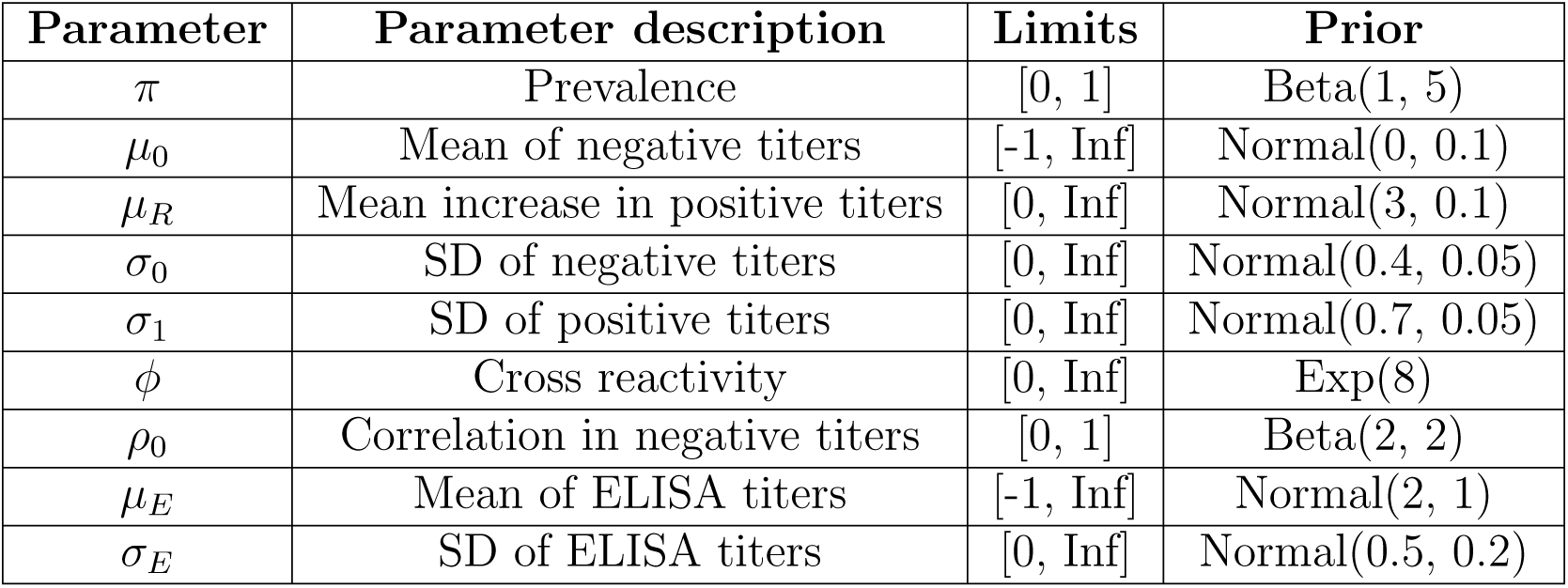
Multivariate mixture model parameter priors. Parameter priors and limits used in model fitting.

#### 1.5 1D mixture model & titer cutoffs

To compare the results of the multivariate Gaussian mixture model framework to traditional approaches, we fit classic single dimension (1D) Gaussian mixture models independently to the antibody titer data from each pathogen. Here, each model fits a two-component Gaussian mixture distribution, with a negative and positive component. The model likelihood is given in equation 20. The same parameter priors for *π*, *µ*_0_, *µ*_1_, *σ*_0_ and *σ*_1_ were used, shown in Table 3, while *ϕ* and *ρ* parameters are not estimated. Cutoffs to classify individuals as positive or negative to each pathogen were calculated as the mean of means of the two Gaussian distributions.

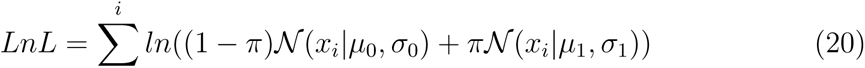

#### 1.6 Catalytic model

For pathogens where age-specific prevalence estimates increased with age indicating endemic transmission dynamics, we fit catalytic models to quantify the annual force of infection (FOI). The FOI, *λ* is the rate at which susceptible individuals become infected each year. We assumed that *λ* is constant over time and age, giving long-term averages of past FOI. The expected proportion of the population positive by age, 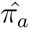, can then be calculated as shown in equation 21. The model was fit in a Bayesian framework using cmdStanR assuming the proportion of positive individuals in each age group to follow a binomial distribution with probability of success equal to 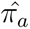. We used a uniform prior between 0 and 1 for *λ* and fit the model with 3 chains of 10,000 warmup iterations plus 10,000 sampling iterations each.

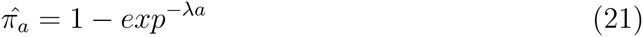

#### 1.7 Multidimensional scaling of antigen-sera relationships

To summarise the antibody cross-reactivity estimates we used multidimensional scaling (MDS) to translate the model median estimates to a 2D map depicting relative antibody-antigen relationships. We used only *µ* estimates from Gaussian components characterizing individuals infected with a single pathogen only, therefore excluding *µ* estimates from infection statuses that were positive to multiple pathogens or no pathogens. We used the Racmacs R package [8] for multidimensional scaling of the model Gaussian *µ* estimates. Scaling the antibody-antigen data to 2 dimensions, we performed 10,000 optimization runs to find the best arrangement of antigens and sera to represent their relative similarities. For each optimization run, points were randomly distributed in 2D space and a limited-memory Broyden–Fletcher–Goldfarb–Shanno (L-BFGS) gradient-based optimization algorithm is applied to find the optimal positions of the points. Antigen-sera distances were extracted from the model fit with the highest likelihood.

#### 1.8 Maximum likelihood phylogeny

We reconstructed a distance-based maximum-likelihood phylogeny of the envelope (E) proteins from each flavivirus considered in this study. We used E protein sequences from reference genomes in NCBI, with accession numbers as follows: WNV (YP 001527880.1), DENV1 (NP 722460.2), DENV2 (NP 739583.2), DENV3 (YP 001531168.2), DENV4 (NP 740317.1), JEV (NP 775666.1), TBEV (NP 775503.1), ZIKV (YP 009227198.1), YFV (NP 740305). As CHIKV is an alphavirus we did not consider it in this analysis. We aligned all protein sequences using MUSCLE [3] and reconstructed the phylogeny using IQ-tree [7], with an LG+G4 substitution model that was automatically selected.

### 2 Supplementary Figures & Tables

**Figure 2.**
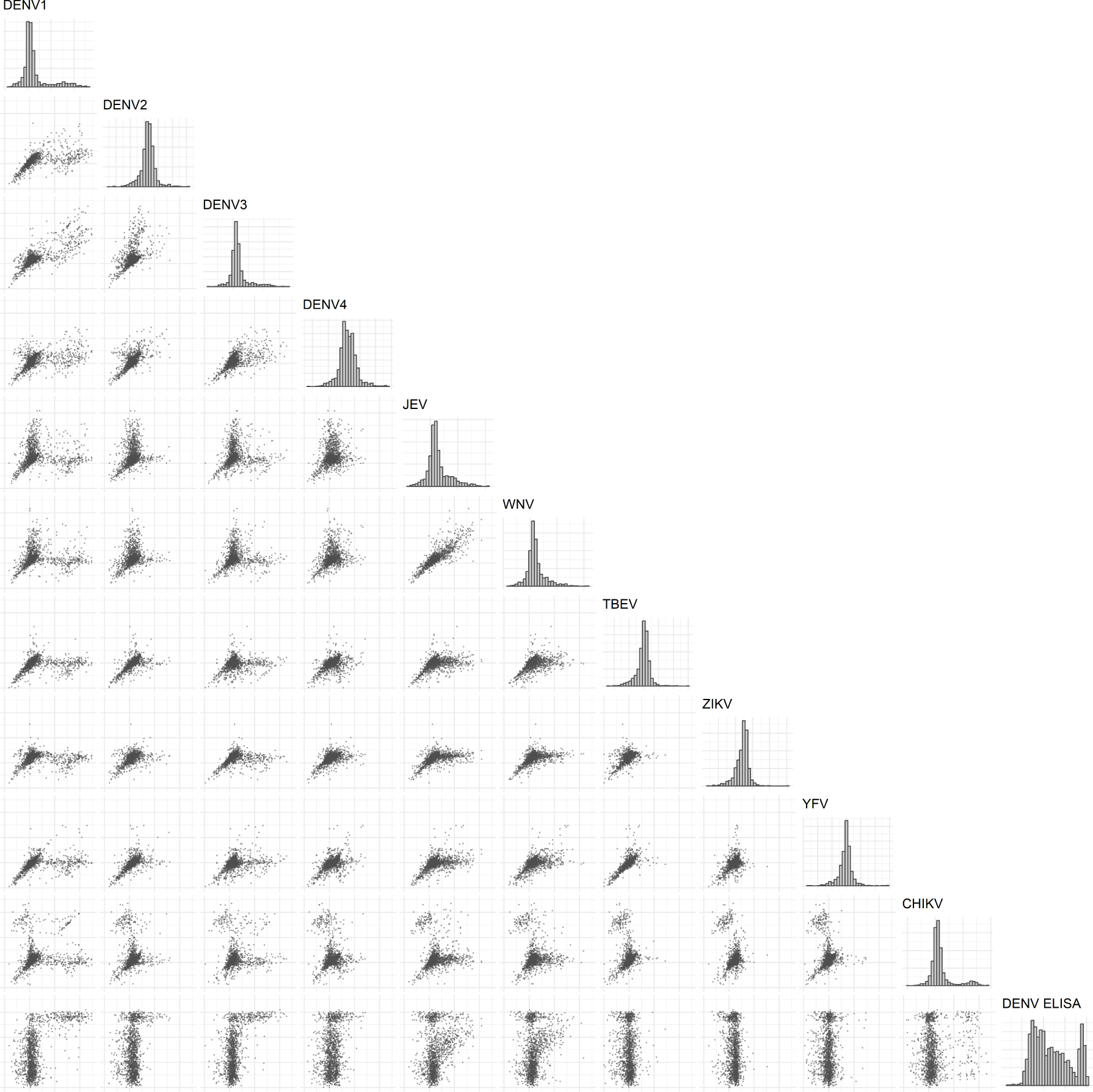
Antibody titer distributions across antigen pairs. Grey bars show the population distribution of measured antibody titers against each antigen, shown on a log relative fluorescence intensity (RFI) scale for the multiplex assay antigens and a log Panbio unit scale for the DENV ELISA assay. Black points in the scatter plots show antibody titer values for each pair of antigens, where the x-axes correspond to antibodies measured against the antigen at the top of each column panels and the y-axes correspond to antibodies measured against the antigen at the right of each row panel.

**Table 4.**
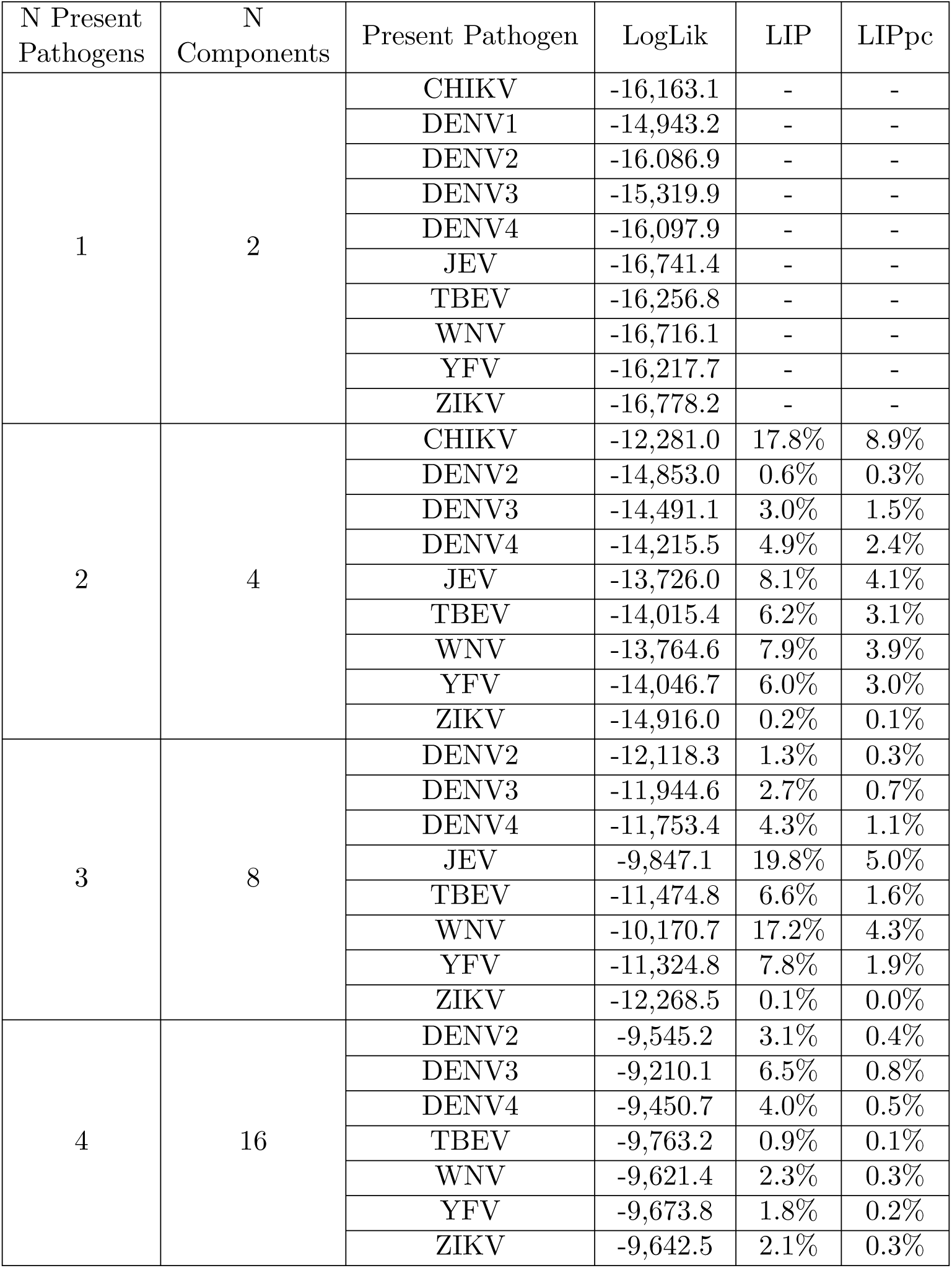
Variable selection process model metrics for base model. Log-likelihood, likelihood increment percentage (LIP) and LIP per component (LIPpc) values from the base model.

| N Present Pathogens | N Components | Present Pathogen | LogLik | LIP | LIPpc |
| --- | --- | --- | --- | --- | --- |
| 1 | 2 | CHIKV | -16,163.1 | - | - |
|  |  | DENV1 | -14,943.2 | - | - |
|  |  | DENV2 | -16,086.9 | - | - |
|  |  | DENV3 | -15,319.9 | - | - |
|  |  | DENV4 | -16,097.9 | - | - |
|  |  | JEV | -16,741.4 | - | - |
|  |  | TBEV | -16,256.8 | - | - |
|  |  | WNV | -16,716.1 | - | - |
|  |  | YFV | -16,217.7 | - | - |
|  |  | ZIKV | -16,778.2 | - | - |
| 2 | 4 | CHIKV | -12,281.0 | 17.8% | 8.9% |
|  |  | DENV2 | -14,853.0 | 0.6% | 0.3% |
|  |  | DENV3 | -14,491.1 | 3.0% | 1.5% |
|  |  | DENV4 | -14,215.5 | 4.9% | 2.4% |
|  |  | JEV | -13,726.0 | 8.1% | 4.1% |
|  |  | TBEV | -14,015.4 | 6.2% | 3.1% |
|  |  | WNV | -13,764.6 | 7.9% | 3.9% |
|  |  | YFV | -14,046.7 | 6.0% | 3.0% |
|  |  | ZIKV | -14,916.0 | 0.2% | 0.1% |
| 3 | 8 | DENV2 | -12,118.3 | 1.3% | 0.3% |
|  |  | DENV3 | -11,944.6 | 2.7% | 0.7% |
|  |  | DENV4 | -11,753.4 | 4.3% | 1.1% |
|  |  | JEV | -9,847.1 | 19.8% | 5.0% |
|  |  | TBEV | -11,474.8 | 6.6% | 1.6% |
|  |  | WNV | -10,170.7 | 17.2% | 4.3% |
|  |  | YFV | -11,324.8 | 7.8% | 1.9% |
|  |  | ZIKV | -12,268.5 | 0.1% | 0.0% |
| 4 | 16 | DENV2 | -9,545.2 | 3.1% | 0.4% |
|  |  | DENV3 | -9,210.1 | 6.5% | 0.8% |
|  |  | DENV4 | -9,450.7 | 4.0% | 0.5% |
|  |  | TBEV | -9,763.2 | 0.9% | 0.1% |
|  |  | WNV | -9,621.4 | 2.3% | 0.3% |
|  |  | YFV | -9,673.8 | 1.8% | 0.2% |
|  |  | ZIKV | -9,642.5 | 2.1% | 0.3% |

**Table 5.** Variable selection process model metrics for location-specific model. Log-likelihood, likelihood increment percentage (LIP) and LIP per component (LIPpc) values from the location-specific model.

| N Present Pathogens | N Components | Present Pathogen | Log-Likelihood | LIP | LIPpc |
| --- | --- | --- | --- | --- | --- |
| 1 | 2 | CHIKV | -16,082.6 | - | - |
|  |  | DENV1 | -14,877.4 | - | - |
|  |  | DENV2 | -16,038.5 | - | - |
|  |  | DENV3 | -15,253.8 | - | - |
|  |  | DENV4 | -16,034.2 | - | - |
|  |  | JEV | -16,733.1 | - | - |
|  |  | TBEV | -16,787.8 | - | - |
|  |  | WNV | -16,705.8 | - | - |
|  |  | YFV | -16,163.1 | - | - |
|  |  | ZIKV | -16,722.5 | - | - |
| 2 | 4 | CHIKV | -12,136.0 | 18.4% | 9.2% |
|  |  | DENV2 | -14,798.2 | 0.5% | 0.3% |
|  |  | DENV3 | -14,414.6 | 3.1% | 1.6% |
|  |  | DENV4 | -14,076.6 | 5.4% | 2.7% |
|  |  | JEV | -13,653.7 | 8.2% | 4.1% |
|  |  | TBEV | -13,883.3 | 6.7% | 3.3% |
|  |  | WNV | -13,694.7 | 7.9% | 4.0% |
|  |  | YFV | -13,905.3 | 6.5% | 3.3% |
|  |  | ZIKV | -14,846.6 | 0.2% | 0.1% |
| 3 | 8 | DENV2 | -11,968.0 | 1.4% | 0.3% |
|  |  | DENV3 | -11,793.9 | 2.8% | 0.7% |
|  |  | DENV4 | -12,023.7 | 0.9% | 0.2% |
|  |  | JEV | -9,690.3 | 20.2% | 5.0% |
|  |  | TBEV | -11,328.5 | 6.6% | 1.7% |
|  |  | WNV | -10,015.9 | 17.5% | 4.4% |
|  |  | YFV | -11,171.5 | 7.9% | 2.0% |
|  |  | ZIKV | -12,123.9 | 0.1% | 0.0% |
| 4 | 16 | DENV2 | -9,379.6 | 3.2% | 0.4% |
|  |  | DENV3 | -9,129.2 | 5.8% | 0.7% |
|  |  | DENV4 | -9,278.0 | 4.3% | 0.5% |
|  |  | TBEV | -9,605.1 | 0.9% | 0.1% |
|  |  | WNV | -9,461.0 | 2.4% | 0.3% |
|  |  | YFV | -9,517.2 | 1.8% | 0.2% |
|  |  | ZIKV | -9,477.3 | 2.2% | 0.3% |

**Table 6.** Variable selection process model metrics for age- and location-specific model. Log-likelihood, likelihood increment percentage (LIP) and LIP per component (LIPpc) values from the age- and location-specific model.

| N Present Pathogens | N Components | Present Pathogen | Log-Likelihood | LIP | LIPpc |
| --- | --- | --- | --- | --- | --- |
| 1 | 2 | CHIKV | -16,088.5 | - | - |
|  |  | DENV1 | -14,877.8 | - | - |
|  |  | DENV2 | -16,037.6 | - | - |
|  |  | DENV3 | -15,253.7 | - | - |
|  |  | DENV4 | -16,036.2 | - | - |
|  |  | JEV | -16,990.6 | - | - |
|  |  | TBEV | -16,203.2 | - | - |
|  |  | WNV | -16,678.5 | - | - |
|  |  | YFV | -16,163.7 | - | - |
|  |  | ZIKV | -17,575.4 | - | - |
| 2 | 4 | CHIKV | -12,138.7 | 18.4% | 9.2% |
|  |  | DENV2 | -14,804.9 | 0.5% | 0.2% |
|  |  | DENV3 | -14,422.4 | 3.1% | 1.5% |
|  |  | DENV4 | -14,077.9 | 5.4% | 2.7% |
|  |  | JEV | -13,589.8 | 8.7% | 4.3% |
|  |  | TBEV | -13,882.6 | 6.7% | 3.3% |
|  |  | WNV | -13,638.6 | 8.3% | 4.2% |
|  |  | YFV | -13,908.7 | 6.5% | 3.3% |
|  |  | ZIKV | -14,863.3 | 0.1% | 0.1% |
| 3 | 8 | DENV2 | -11,971.8 | 1.4% | 0.3% |
|  |  | DENV3 | -11,797.3 | 2.8% | 0.7% |
|  |  | DENV4 | -12,031.3 | 0.9% | 0.2% |
|  |  | JEV | -9,638.0 | 20.6% | 5.2% |
|  |  | TBEV | -11,314.5 | 6.8% | 1.7% |
|  |  | WNV | -9,960.2 | 17.9% | 4.5% |
|  |  | YFV | -11,149.8 | 8.1% | 2.0% |
|  |  | ZIKV | -12,108.7 | 0.2% | 0.1% |
| 4 | 16 | DENV2 | -9,330.2 | 3.2% | 0.4% |
|  |  | DENV3 | -9,079.4 | 5.8% | 0.7% |
|  |  | DENV4 | -9,229.0 | 4.2% | 0.5% |
|  |  | TBEV | -9,565.2 | 0.8% | 0.1% |
|  |  | WNV | -9,404.0 | 2.4% | 0.3% |
|  |  | YFV | -9,471.1 | 1.7% | 0.2% |
|  |  | ZIKV | -9,430.2 | 2.2% | 0.3% |
| 5 | 32 | DENV2 | -8,720.5 | 4.0% | 0.2% |
|  |  | DENV4 | -8,474.5 | 6.7% | 0.4% |
|  |  | TBEV | -8,653.4 | 4.7% | 0.3% |
|  |  | WNV | -8,730.9 | 3.8% | 0.2% |
|  |  | YFV | -8,880.1 | 2.2% | 0.1% |
|  |  | ZIKV | -8,773.1 | 3.4% | 0.2% |
| 6 | 64 | DENV2 | -8,130.7 | 4.1% | 0.1% |
|  |  | TBEV | -8,321.9 | 1.8% | 0.1% |
|  |  | WNV | -8,004.6 | 5.5% | 0.2% |
|  |  | YFV | -8,186.3 | 3.4% | 0.1% |
|  |  | ZIKV | -8,397.1 | 0.9% | 0.0% |

**Figure 3.**
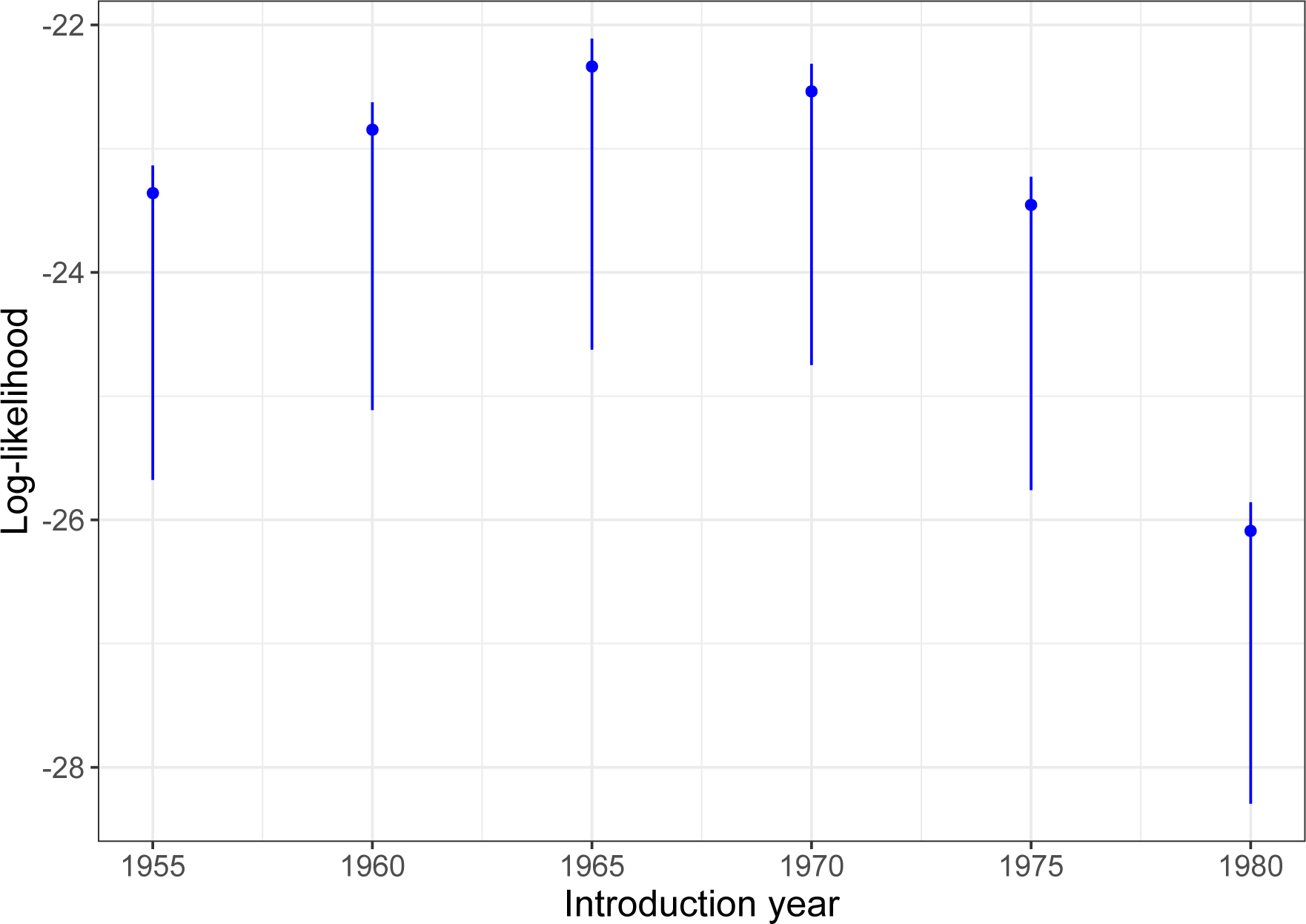
Catalytic model log-likelihood by assumed JEV introduction time. Blue points and lines show the model median and 95%CrI log-likelihood estimates by assumed introduction time of JEV.

**Figure 4.**
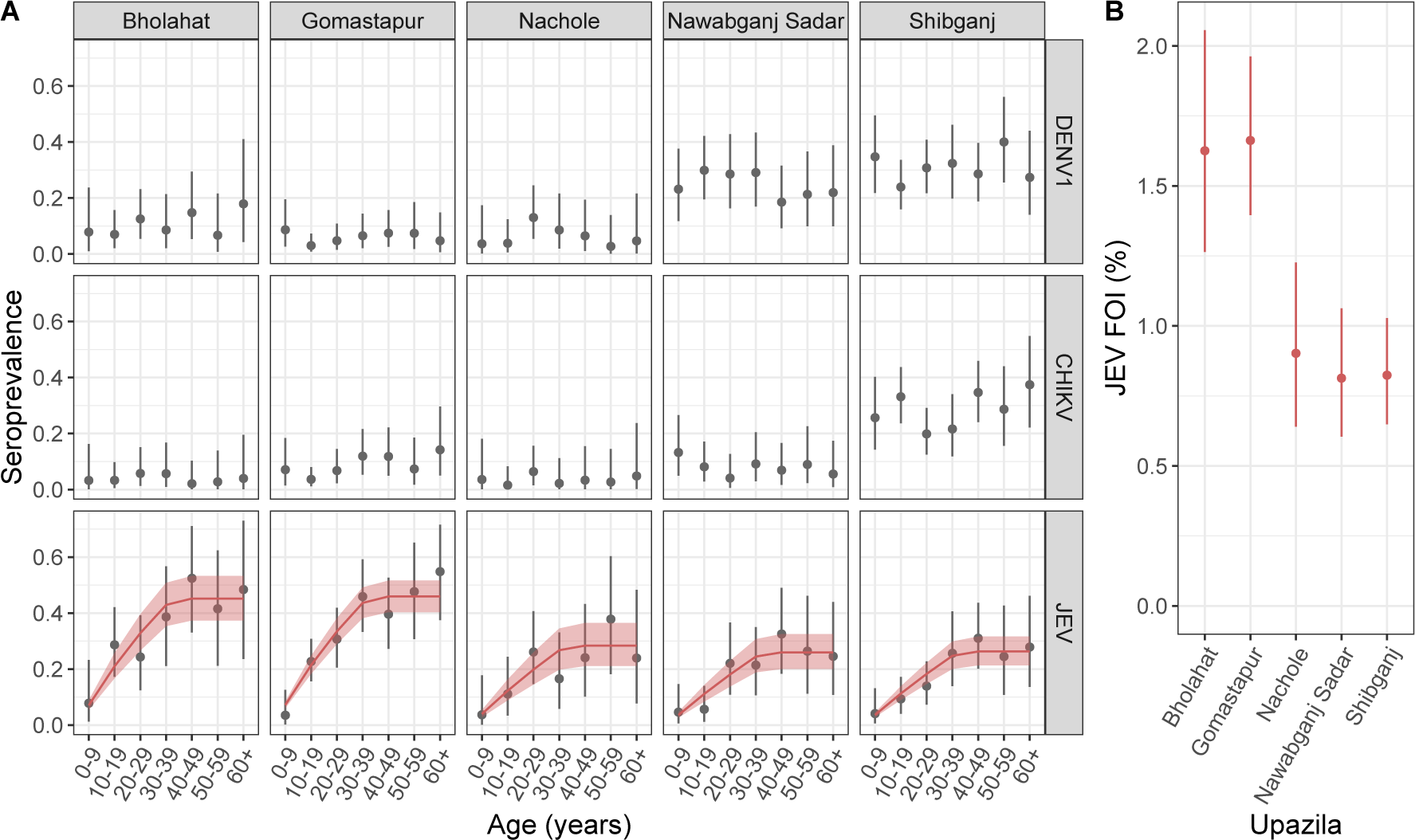
Infection prevalence estimates by pathogen, location and age. Panel A shows estimates of infection prevalence by pathogen, upazila and age group, where grey points and lines indicate median and 95%CrI estimates. Red lines and shaded ribbons show the fit of catalytic models to the infection prevalence estimates, assuming constant endemic JEV transmission since 1977 [5]. Panel B shows the estimates of JEV force of infection (FOI) by upazila.

**Figure 5.**
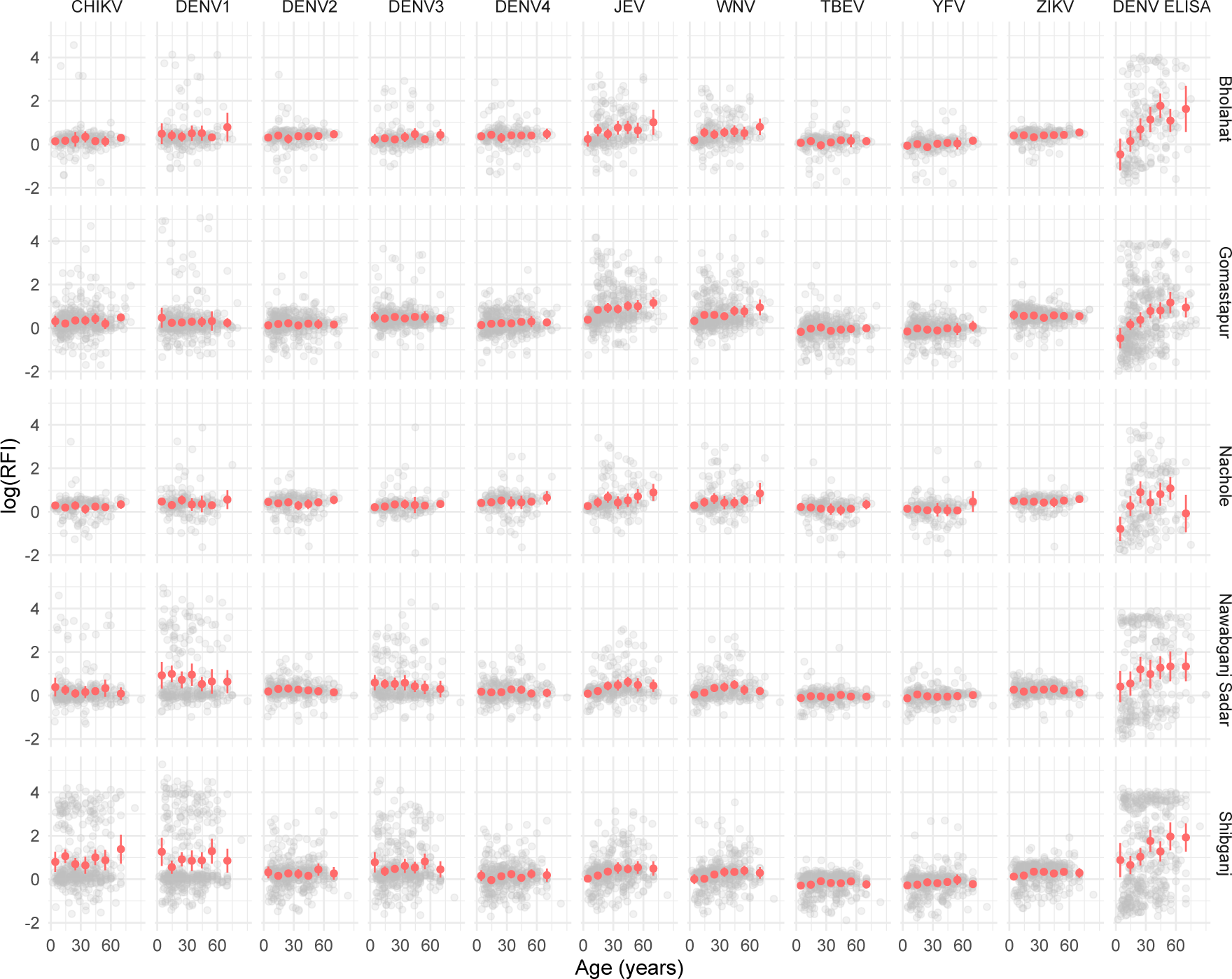
RFI by pathogen, location and age. Grey points show the log relative fluorescence intensity (RFI) values for each individual by age shown on the x-axis, antigen (column panels), and sub-district (row panels). Red points and lines show the population mean and 95% confidence interval estimates of log RFI by age group. Individuals were grouped by 10-year age groups from 0-60 years and individuals aged 60+ were aggregated to a single age group. Red points and lines are plotted at the respective age group mid-points.

**Figure 6.**
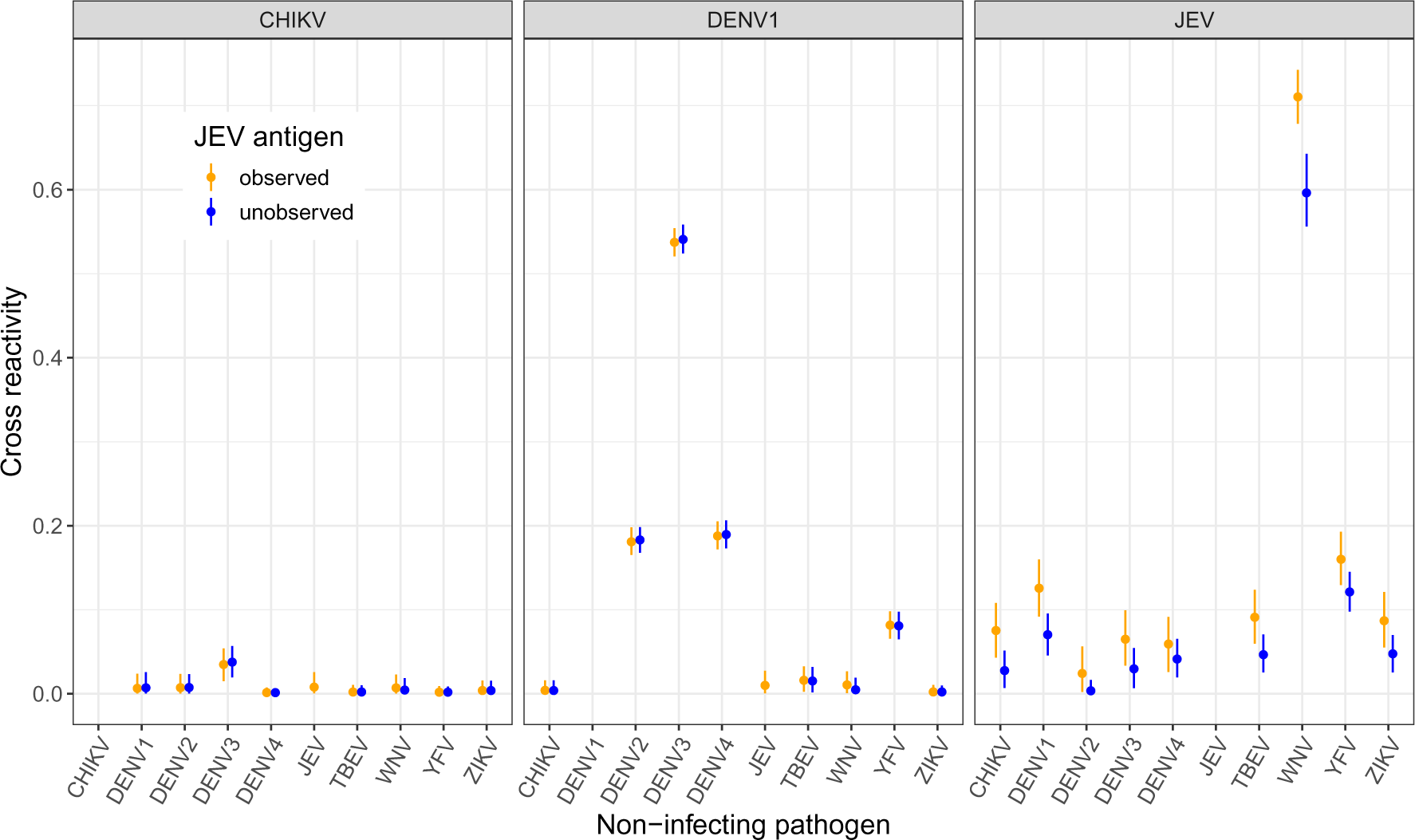
Reconstructed cross reactivity estimates when JEV is unobserved. Cross reactivity estimates, defined as relative titer increases to the non-infecting pathogen compared to the infecting pathogen, model estimates. Points and lines indicate median and 95%CrI model estimates. Estimates colored in orange represent results where JEV titers are included in model fitting and JEV is assumed to be present. Blue colored estimates represent results of a model where the JEV titers were excluded from fitting but parameters for an unobserved “pathogen X” were inferred. Each panel represents the infecting pathogen.

**Figure 7.**
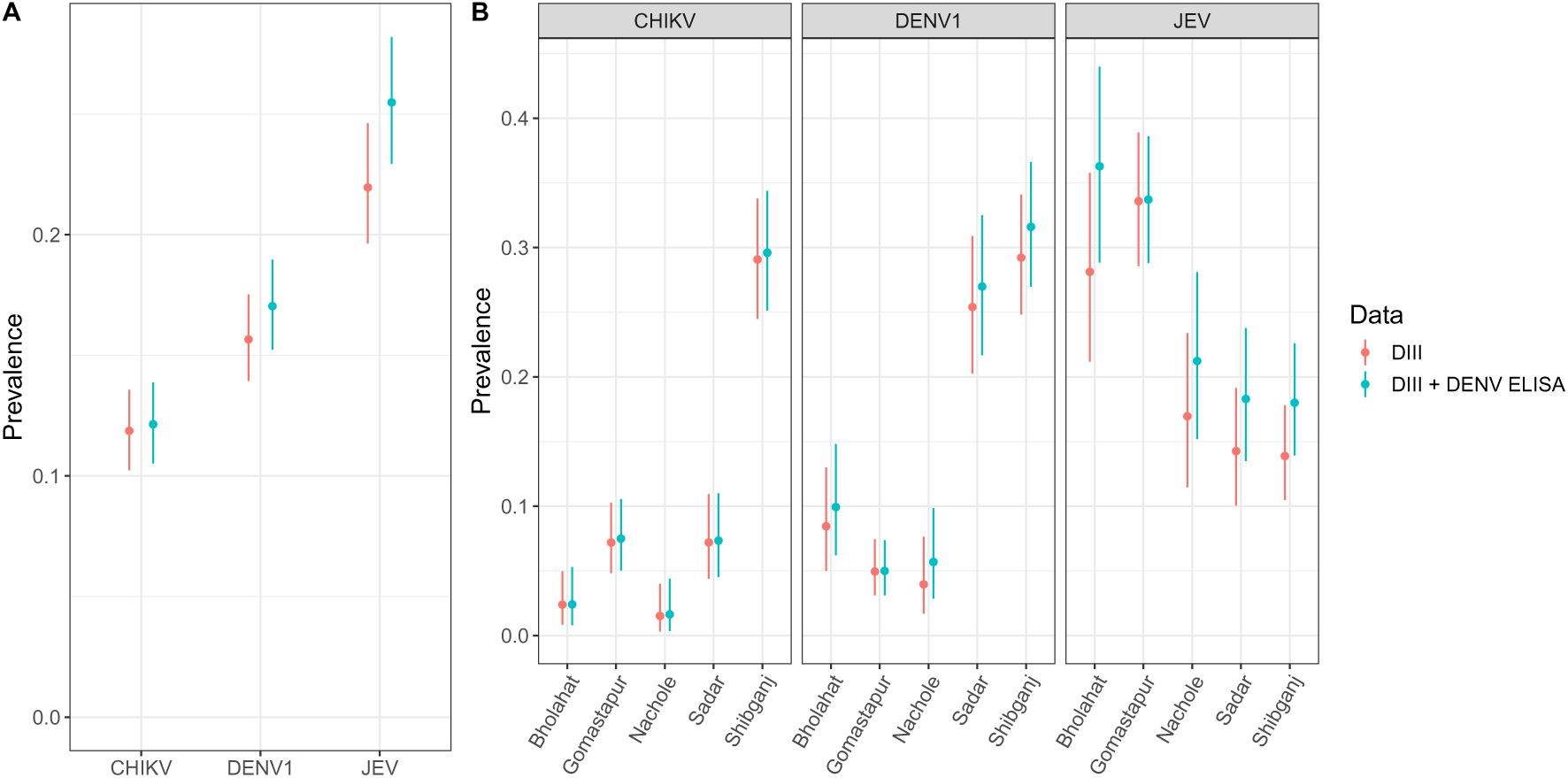
Infection prevalence estimates by assay data. Panel A shows the prevalence of infection per pathogen estimated by the location and age model by assay data for the district of Chapai Nawabganj. Panel B shows the prevalence of infection estimates by pathogen and sub-district depending on which data was included in the model. Points and lines show the median and 95%CrI model estimates.

### 3 Simulation testing

#### 3.1 Methods

To assess the performance of the model and understand its applications we conduct simulation testing across a range of scenarios. Parameter values for simulation were randomly drawn using Latin Hypercube sampling to ensure the approximately even distribution of parameter combinations across simulations. Parameter values for simulation were sampled from uniform distributions, with the ranges of values considered shown in Table 7. The ranges of possible parameter values were selected to approximately reflect antibody titer concentrations on a log relative fluorescence intensity (RFI) scale, where median fluorescence intensity (MFI) values are divided by individual-level background control values before taking the natural logarithm. The model was fit using uninformative priors on the infection prevalence and cross-reactivity parameters, while weakly informative priors were placed on the Gaussian mean and standard deviation parameters (Table 7).

**Table 7.** Parameter ranges and priors for simulation study. Ranges of possible parameter values considered for simulation using Latin Hypercube sampling. Parameters relating to antibody titers are considered on a log scale of the relative fluorescent intensity (RFI).

| Parameter | Parameter description | Simulation Limits | Prior |
| --- | --- | --- | --- |
| $\pi$ | Infection prevalence | [0, 1] | Beta(1, 1) |
| $\mu_0$ | Mean of negative titers | [-0.5, 0.5] | Normal(0, 0.2) |
| $\mu_1$ | Mean increase in positive titers | [1, 3.5] | Normal(2.5, 0.5) |
| $\sigma_0$ | SD of negative titers | [0.25, 0.5] | Normal(0.4, 0.1) |
| $\sigma_1$ | SD of positive titers | [0.5, 0.75] | Normal(0.6, 0.1) |
| $\phi$ | Cross-reactivity | $[0, 1]$ | Beta(1, 1) |
| $\rho$ | Correlation coefficients | $[0, 1]$ | Beta(1, 1) |

We first assess the performance of the model on a 2 pathogen system, simulating antibody titers against 2 related pathogens and assessing the models ability to reconstruct parameter values from 50 simulated datasets. We further assess model performance with an increasing number of pathogens, considering systems of 3, 5 and 7 related pathogens. For all simulations with *≤* 3 pathogens, we assume the Gaussian standard deviation parameters, *σ*_0_ and *σ*_1_ to be independent for each pathogen. For simulations with *>* 3 pathogens, however, we assume constant *σ*_0_ and *σ*_1_ across pathogens which may be a reasonable assumption when antibodies against each antigen are measured using the same assay. In each case we generate 50 simulated datasets of 1,500 individuals and fit the model to each of the simulations. All models were fit with 2 chains for 3,000 iterations in addition to 3,000 warm-up samples. Model convergence was assessed by visual inspection of chain mixing and by R-hat convergence diagnostic across all parameters. When comparing model performance we assess differences in deviance information criterion (DIC), widely applicable information criteria (WAIC) and leave-one-out expected log pointwise predictive density (ELPD LOO) metrics. When comparing the performance of models with varying numbers of Gaussian components we additionally use LIPpc and LIPpp metrics. Similar to the LIPpc metric described in section 1.4.1, the likelihood increment percentage per parameter (LIPpp) penalizes the likelihood increment by the additional number of parameters used in the more complex model.

##### 3.1.1 Heterogeneity in infection prevalence

In this section we consider a spatially-stratified 2 pathogen system with sampling conducted in 3 locations with varying levels of pathogen-specific infection prevalence across locations. We assume a total sample size of 1,500 individuals across the 3 locations, with 500 individuals from each. As before, Latin Hypercube sampling was used to randomly draw parameter values for 50 simulations with the same possible parameter ranges shown in Table 7. Only infection prevalence parameters were varied by location and all other parameters were assumed to be constant across locations. To each simulation we fit two model versions; a base model version where no location information is used and a single value of pathogen-specific infection prevalence is estimated for the entire population, and a second location-specific version where pathogen-specific infection prevalence is allowed to vary across the three locations.

##### 3.1.2 1D mixtures

To understand the biases that can occur when antibody cross-reactivity is present but not accounted for, we fit classic single-dimension mixture models independently to antibody titer data for each pathogen using the 2 pathogen simulations. Here, each model fits a two-component Gaussian mixture distribution, with a negative and positive component as described above in section 1.5.

##### 3.1.3 Absent pathogens

To understand the ability of the model to differentiate between pathogen presence and absence in systems of related cross-reacting pathogens, we conduct further simulations. Using the same 2 and 3 pathogen simulations from the previous section, we set the infection prevalence of one pathogen to zero and re-simulate the antibody titer data. We then fit the same base model to these new simulations as well as an “absent model” that assumes the absence of the truly absent pathogen.

#### 3.2 Results

##### 3.2.1 Model performance for two pathogen systems

We found high consistency between true and median estimates of infection prevalence with an adjusted *R*^2^ of 0.99, shown in Figure 8. True values of prevalence, *π*, were within model 95%CrI estimates in 98% of simulations (49/50) and in 99% of prevalence estimates (99/100) (Figure 8). For the cross-reactivity parameters, *ϕ*, high consistency was observed between true and estimated parameter values with an adjusted *R*^2^ of 0.83 (Figure 8). 94% of estimates (94/100) contained the true values of *ϕ* within the 95%CrIs, while 90% (45/50) of simulations accurately estimated both *ϕ* parameters. Only one simulation produced non-accurate estimates of *ϕ* for both parameters, the same simulation that did not accurately estimate the prevalence of pathogen A (Simulation 40). In this simulation, individuals infected with only pathogen A had higher titers against pathogen B than individuals truly infected with pathogen B, causing unidentifiability of the Gaussian components. We also note that simulations with inaccurate estimates of *ϕ* and/or *ρ* tended to occur for simulations with prevalence values close to 0% or 100%. In these simulations, 1 or more Gaussian component could not be characterised due to a limited number of data points in that component.

**Figure 8.**
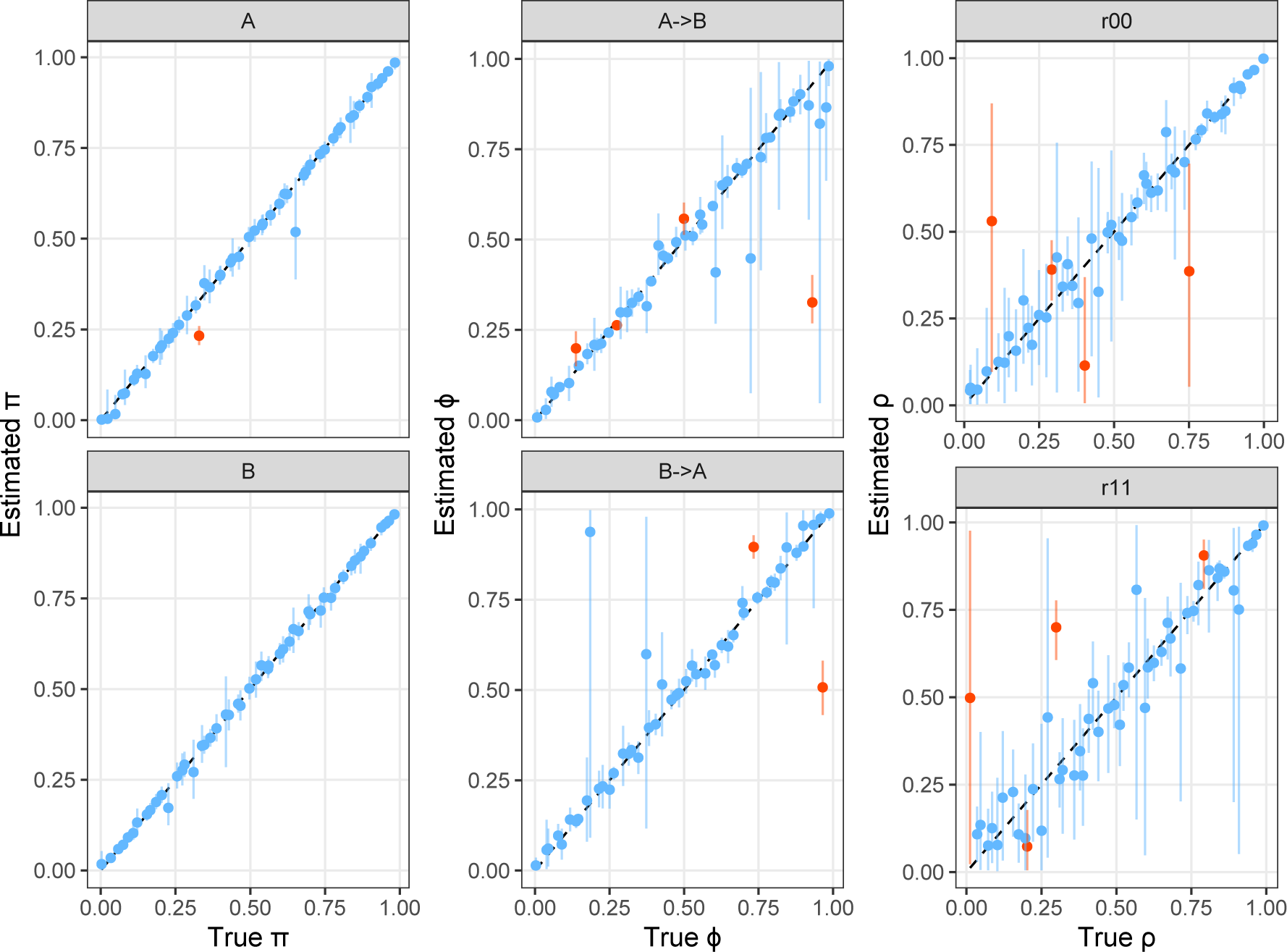
Estimated vs true infection prevalence and cross-reactivity parameters in 2D simulations. Coloured points and lines show the median and 95%CrI model estimates of infection prevalence, cross-reactivity and correlation parameters, compared to the true values in 50 simulations. The black dashed line indicates where estimated values would equal the true values. Blue points and lines highlight the model estimates where the true value was contained within the 95%CrI and orange highlight those that did not contain the true value within estimated 95%CrI. Panels A-*>*B and B-*>*A represent the estimates of relative titer increase from A to B and B to A, respectively. Panels r00 and r11 represent the correlation in the Gaussian distribution for individuals negative to both pathogens, *ρ*_00_ and positive to both pathogens, *ρ*_11_, respectively.

A high level of consistency between true and estimated correlation parameters, *ρ*_00_ and *ρ*_11_, was also observed with adjusted *R*^2^ values of 0.88 and 0.84, respectively (Figure 8). True parameter values were within the estimated 95%CrIs for 92% and 92% of estimates for *ρ*_00_ and *ρ*_11_. The means and standard deviations of the Gaussian components were also well identified by the model, with adjusted *R*^2^ values of 0.96 and 0.97 between true and estimated values for *µ*_0_ and *µ*_1_, and of 0.84 and 0.78 for *σ*_0_ and *σ*_1_ parameters respectively, shown in Figure 9. True parameter values were contained within the 95%CrIs for 96% and 87% of estimates for *µ*_0_ and *µ*_1_ parameters and in 94% and 96% of estimates for *σ*_0_ and *σ*_1_ parameters (Figure 9). Model fits to the the simulated data are shown in Figure 10. We note that the model demonstrates high levels of performance even when large overlap of negative and positive titers result in unimodal titer distributions (Figure 10).

**Figure 9.**
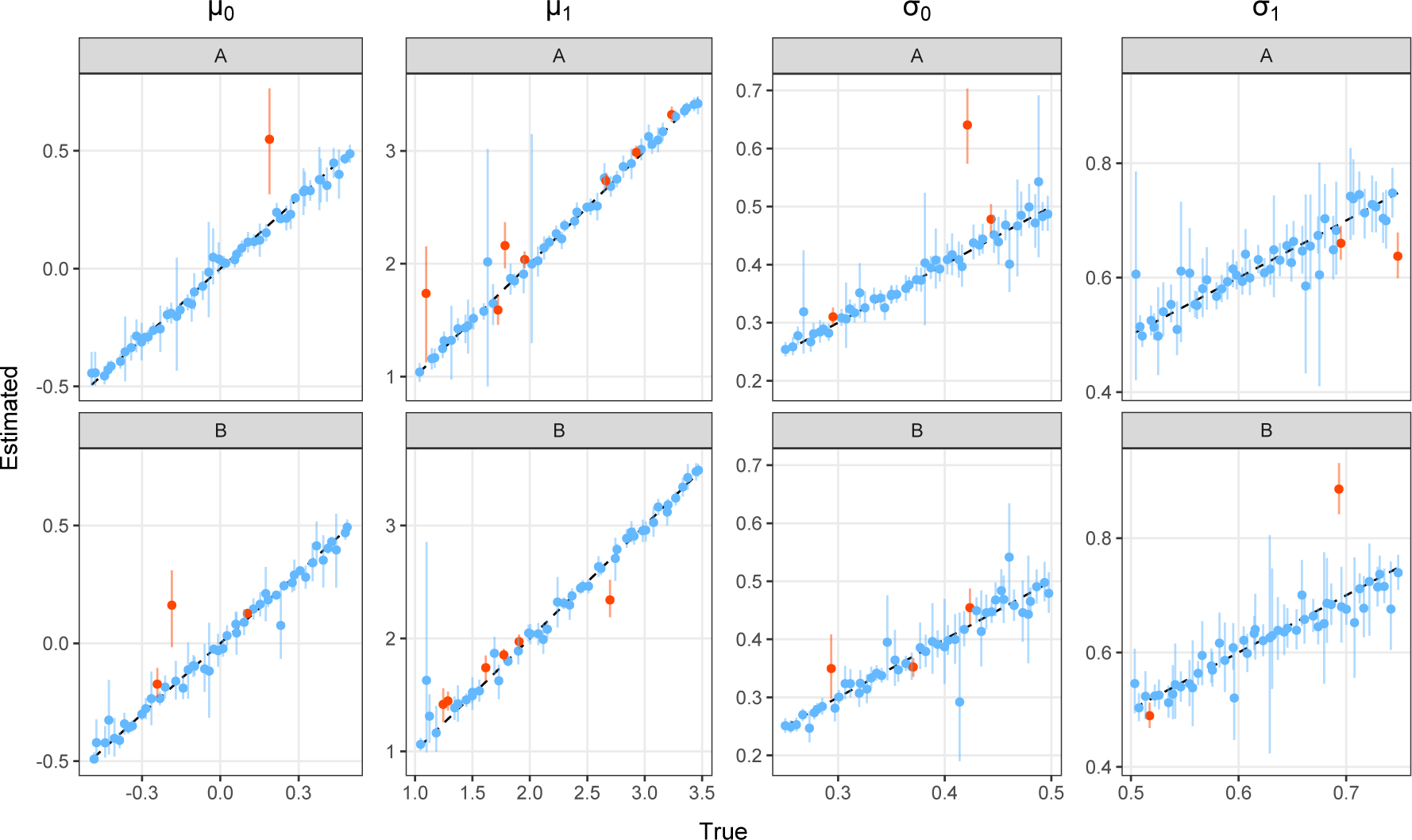
Estimated vs true Gaussian mean and standard deviation parameters in 2D simulations. Coloured points and lines show the median and 95%CrI model estimates of the *µ* and *σ* parameters, compared to the true values in 50 simulations. Each column of panels corresponds to a different parameter and each row represents estimates for pathogen A or pathogen B. The black dashed line indicates where estimated values would equal the true values. Blue points and lines highlight the model estimates where the true value was contained within the 95%CrI and orange highlight those that did not contain the true value within estimated 95%CrI.

**Figure 10.**
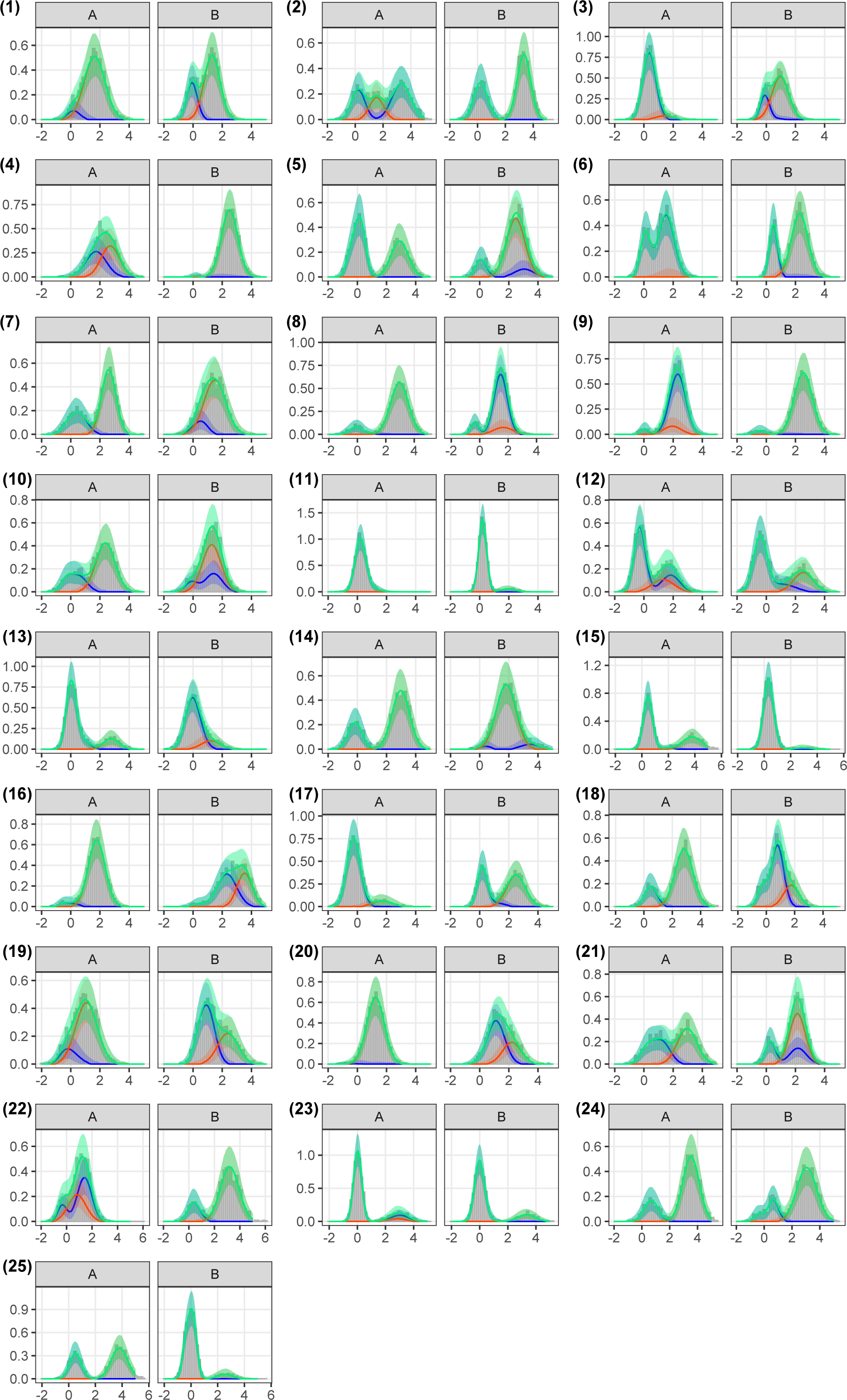
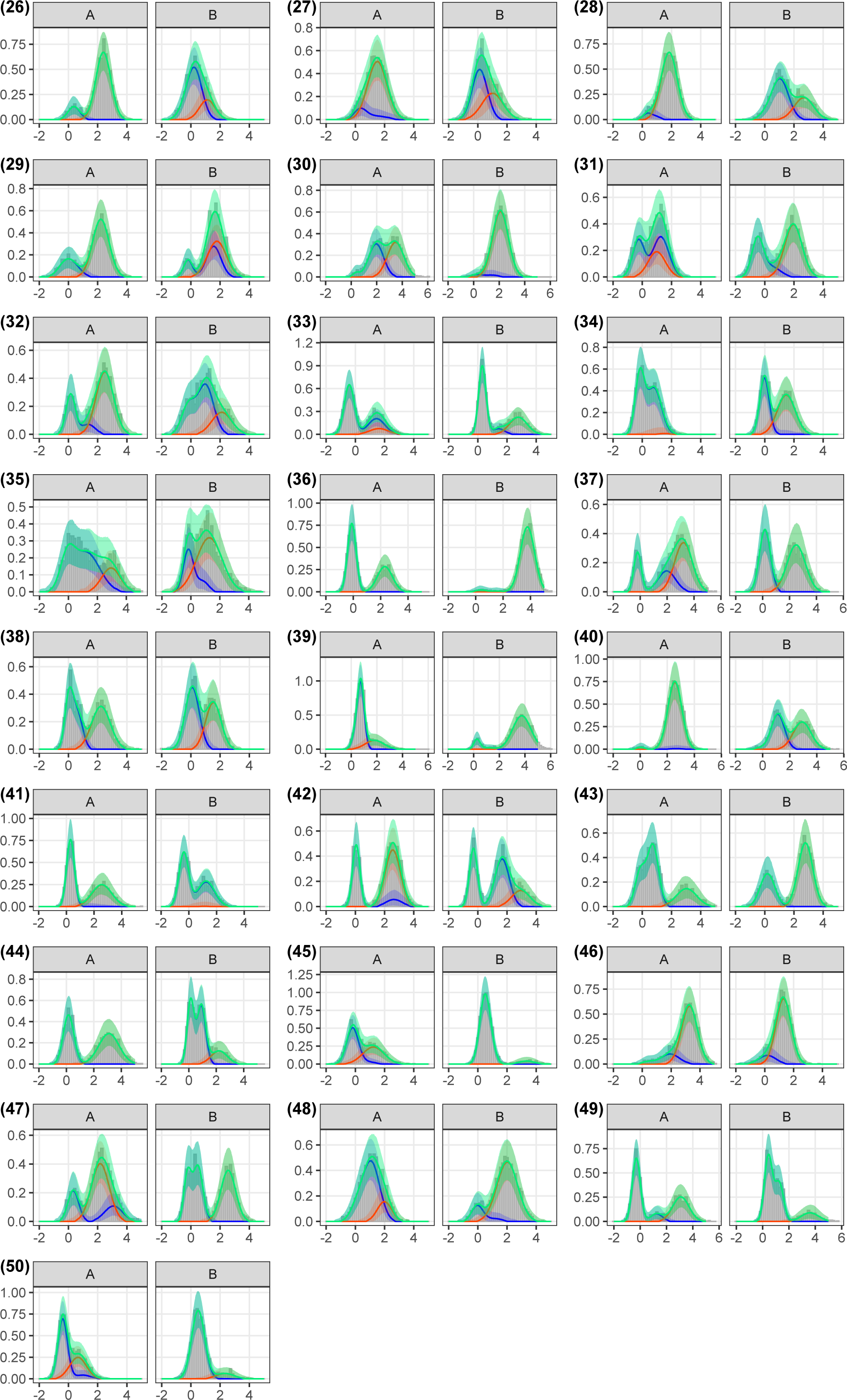
Model fits to 2D simulations. Grey bars show the density distribution of simulated antibody titers for each of the 50 simulations, for pathogens A and B. The lines and shaded ribbons show the median and 95%CrI model reconstructed titer distributions. Blue and orange indicates the reconstructed distribution of negative and positive titers to each pathogen respectively, while green shows the overall reconstructed titer distribution (combined across all components).

##### 3.2.2 Model performance with increasing pathogen numbers

We fit the model to simulated data with an increasing number of pathogens, considering systems of 3, 5 and 7 related pathogens. For each of these considered systems we simulated 50 antibody titer datasets and fit the model to each. Due to non-convergence of some models, 0, 1 and 8 model results were removed from the following summary results from the 3D, 5D and 7D simulations respectively. High consistency was observed between true and estimated prevalence with adjusted *R*^2^ values of 0.95, 1.00 and 1.00 for 3D, 5D and 7D simulations as shown in Figure 11, panel (a). True infection prevalence values were contained within 95%CrIs in 93% of estimates (140/150 estimates) for 3D simulations, 98% of estimates (240/245) for 5D simulations and 99% of estimates (293/294) for 7D simulations (Figure 11 panel (a)). It is worth noting that the 3D simulations assumed independent *σ*_0_ and *σ*_1_ parameters per pathogen in line with the potential use of different assays for each pathogen. In contrast, for 5D and 7D simulations and models we assumed constant *σ*_0_ and constant *σ*_1_ across pathogens.

For cross-reactivity parameters, *ϕ*, the model showed good performance with adjusted *R*^2^ values between estimated and true parameter values of 0.94, 0.96 and 0.98 for 3D, 5D and 7D simulations respectively, shown in Figure 11 panel (b). True values were contained within estimated 95%CrIs for 90% of estimates (270/300) in 3D simulations, 95% of estimates (928/980) in 5D simulations and 92% of estimates (1631/1764) in 7D simulations (Figure 11 panel (b) and Figure 12). Adjusted *R*^2^ values between estimated and true values of *ρ*_00_ parameters were 0.95, 0.99 and 1.00 for 3D, 5D and 7D simulations respectively, with coverage of true values contained within 95%CrIs in 88% (44/50), 92% (45/49) and 93% (39/42) of simulations, shown in Figure 13 and Figure 12. For *ρ*_11_ parameter values, adjusted *R*^2^ values between estimated and true were 0.88, 1.00 and 1.00 for 3D, 5D and 7D simulations with coverage of true values for 86% (43/50), 94% (46/49) and 95% (40/42) of simulations, shown in Figure 13 and Figure 12.

Estimates of *µ*_0_ contained true parameter values within CrIs for 93% (139/150), 95% (233/245) and 91% (268/294) estimates for 3D, 5D and 7D simulations, respectively, shown in Figure 12 and Figure 13. For *µ*_1_ parameters, estimates contained true values in 90% (135/150), 97% (238/245) and 90% (266/294) of estimates for 3D, 5D and 7D simulations (Figure 12). Estimates of *σ*_0_ contained true parameter values within CrIs for 91% (136/150), 79% (194/245) and 100% (294/294) estimates for 3D, 5D and 7D simulations. While for *σ*_1_, estimates contained true parameter values within CrIs for 88% (132/150), 87% (214/245) and 98% (287/294) estimates for 3D, 5D and 7D simulations. Model fits to the 3 pathogen simulations are shown in Figure 14.

**Figure 11.**
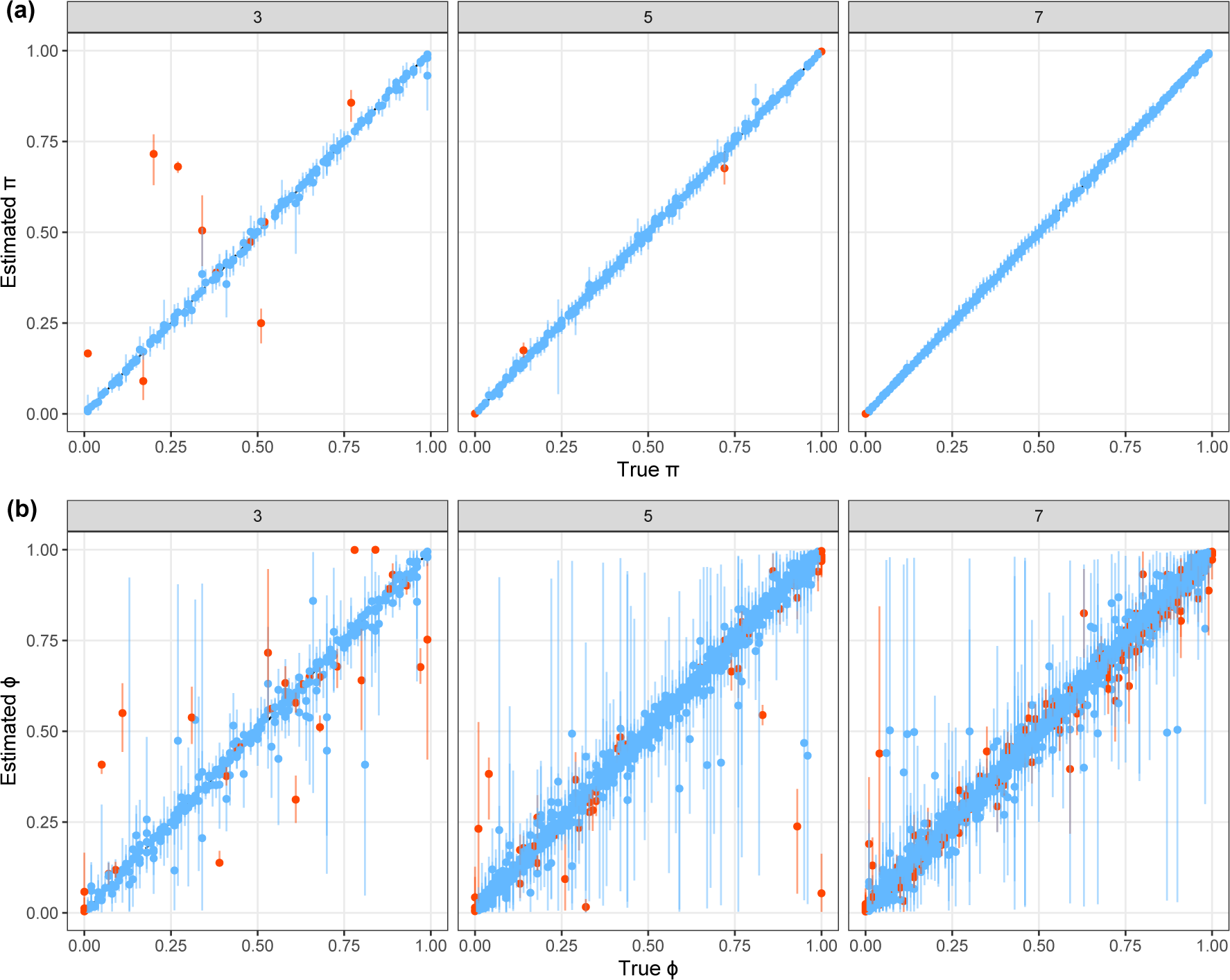
Estimated vs true infection prevalence and cross-reactivity parameters in 3D, 5D and 7D simulations. Coloured points and lines show the median and 95%CrI model estimates of (a) prevalence, *π*, and (b) cross-reactivity, *ϕ*, parameters compared to the true values in 50 simulations. Panels indicate the number of pathogens (3, 5 or 7). The black dashed line indicates where estimated values would equal the true values. Blue points and lines highlight the model estimates where the true value was contained within the 95%CrI and orange highlight those that did not contain the true value within estimated 95%CrI.

**Figure 12.**
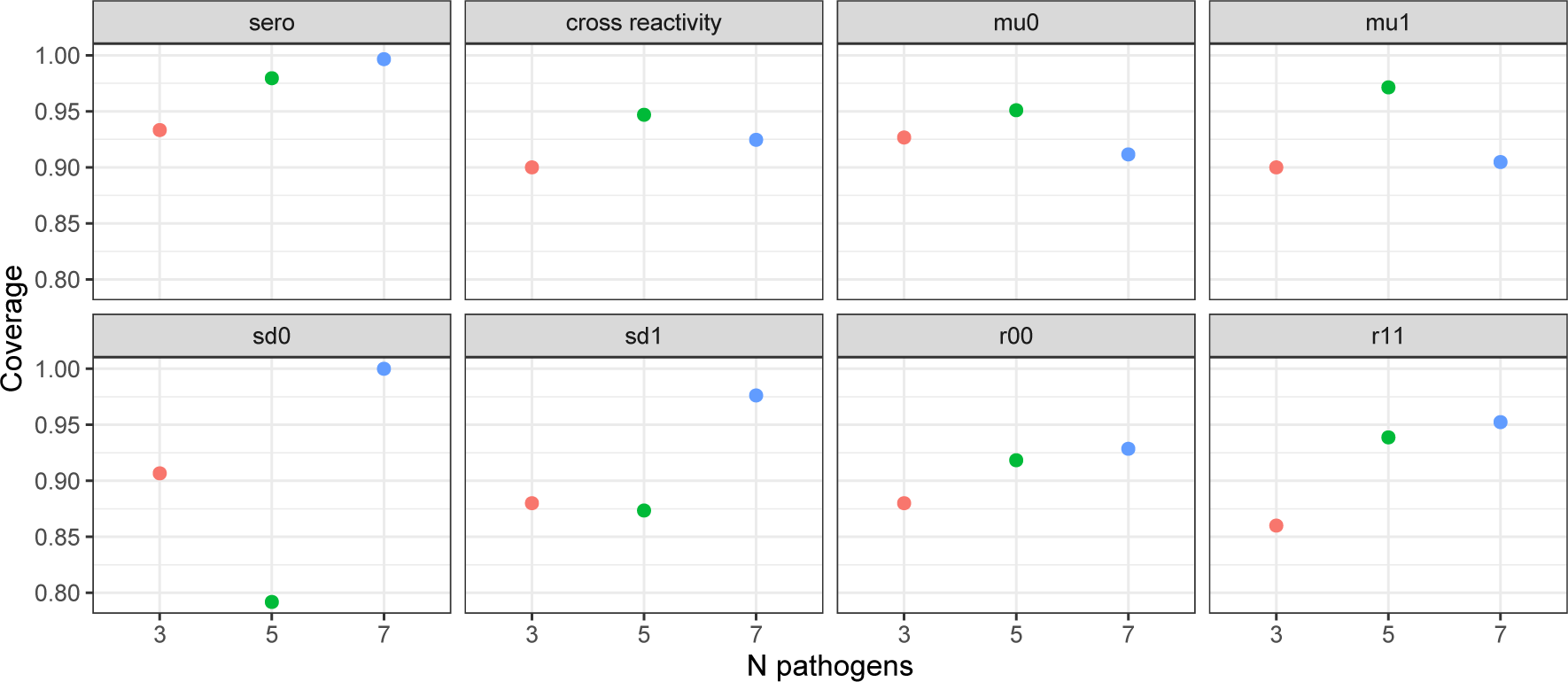
Coverage of true parameter values within credible intervals in 3D, 5D and 7D simulations. Coloured points show the proportion of estimated parameter values that contained the true value within the 95%CrI estimates for 3D, 5D and 7D simulations. Each panel represents a different parameter - infection prevalence (*π*), cross reactivity (*ϕ*), Gaussian means (*µ*_0_ and *µ*_1_), Gaussian standard deviations (*σ*_0_ and *σ*_1_), and correlation parameters (*ρ*_00_ and *ρ*_11_).

**Figure 13.**
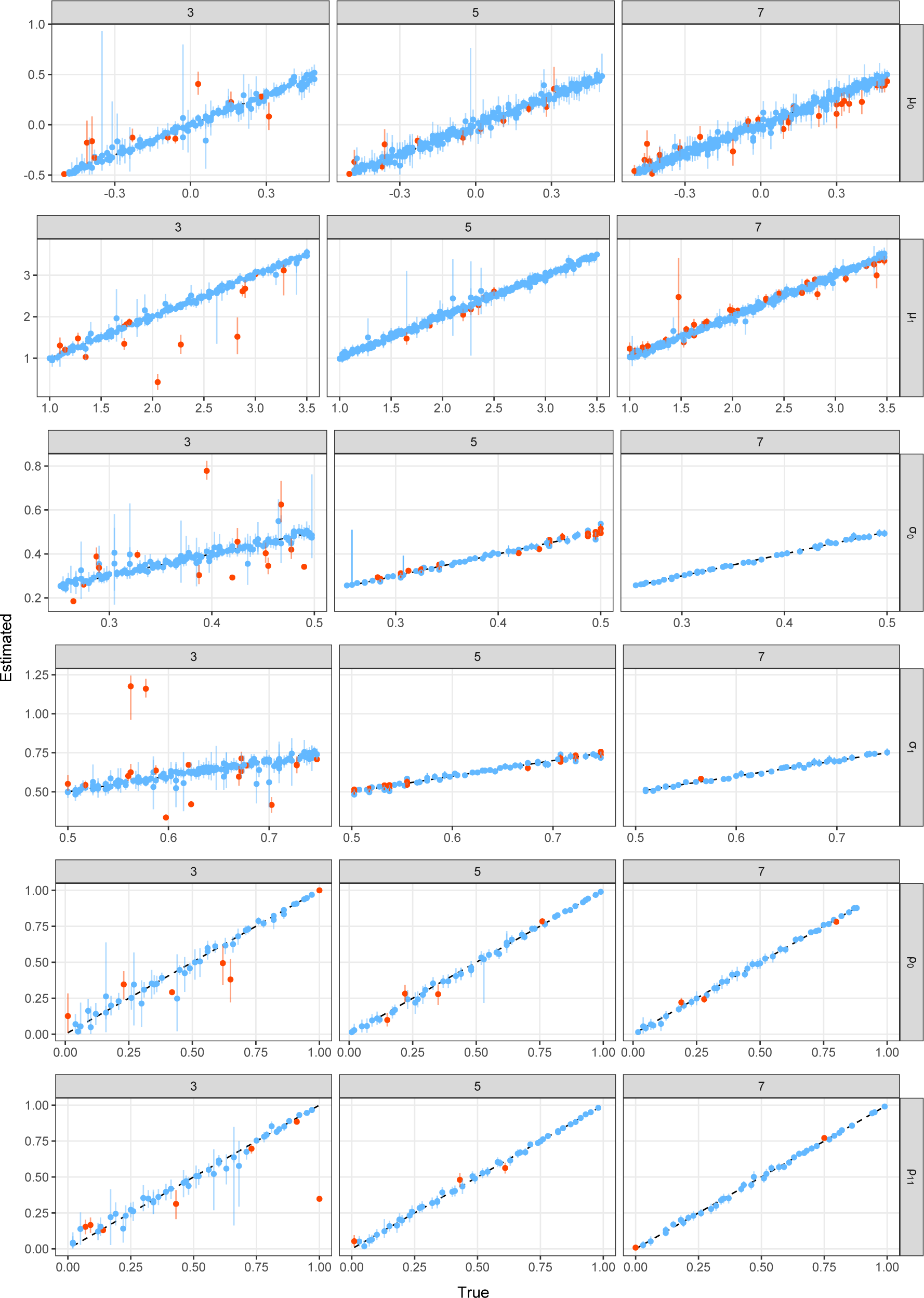
Estimated vs true Gaussian parameters in 3D, 5D and 7D simulations. Coloured points and lines show the median and 95%CrI model parameter estimates compared to the true values in 50 simulations. Columns indicate the number of pathogens (3, 5 or 7), while rows represent each parameter. The black dashed line indicates where estimated values would equal true values. Blue points and lines highlight the model estimates where the true value was contained within the 95%CrI and orange highlight those that did not contain the true value within estimated 95%CrI.

**Figure 14.**
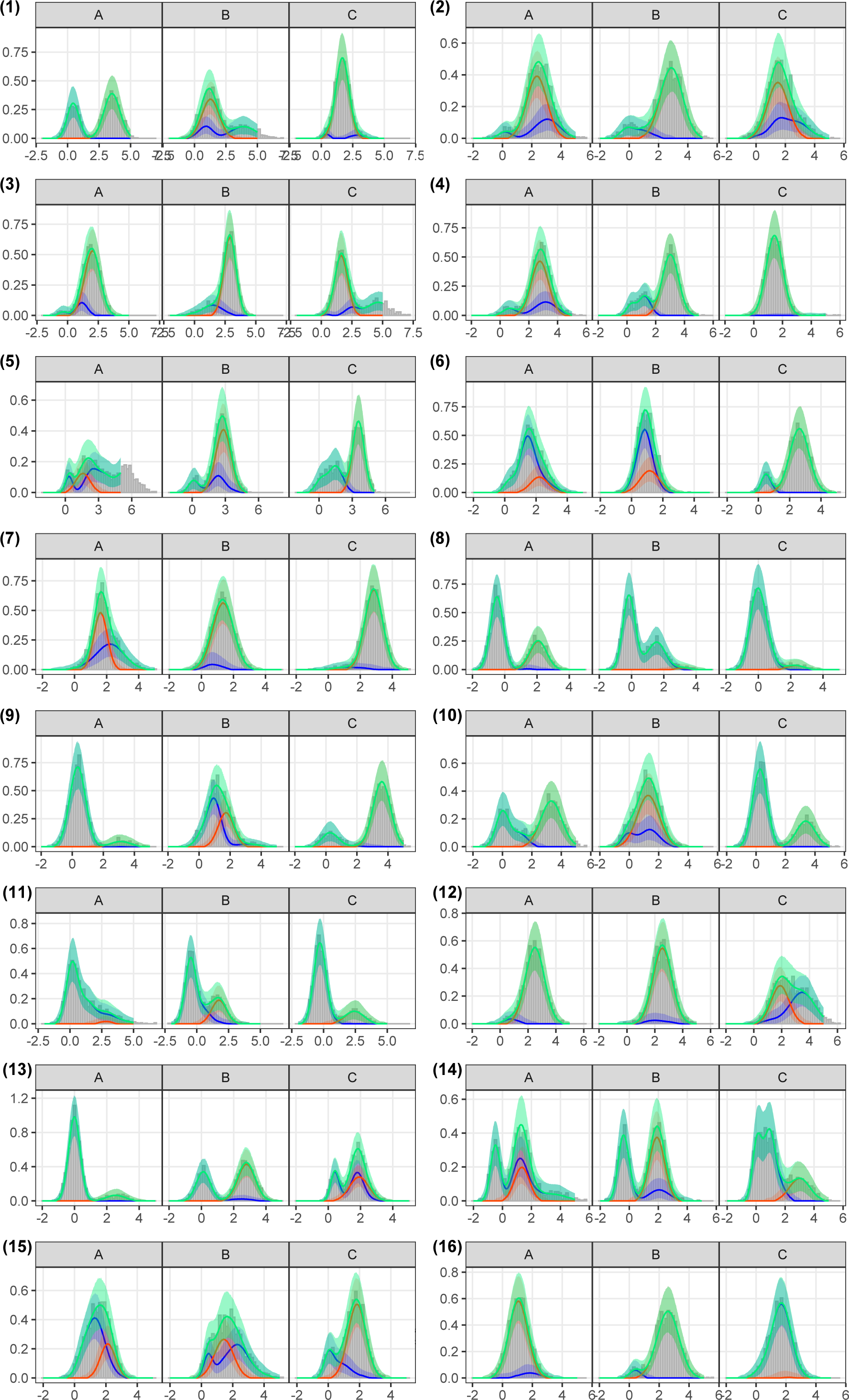
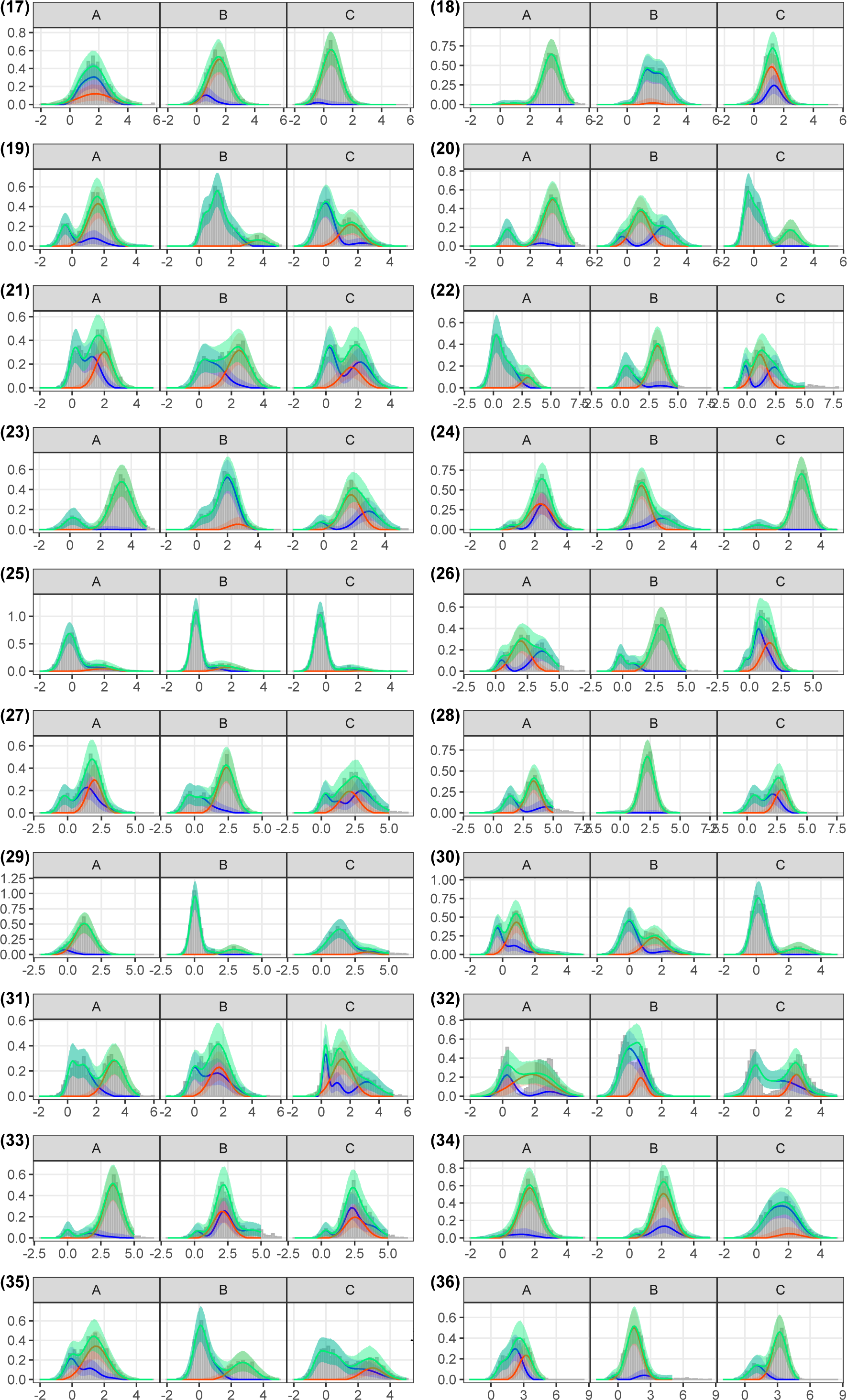
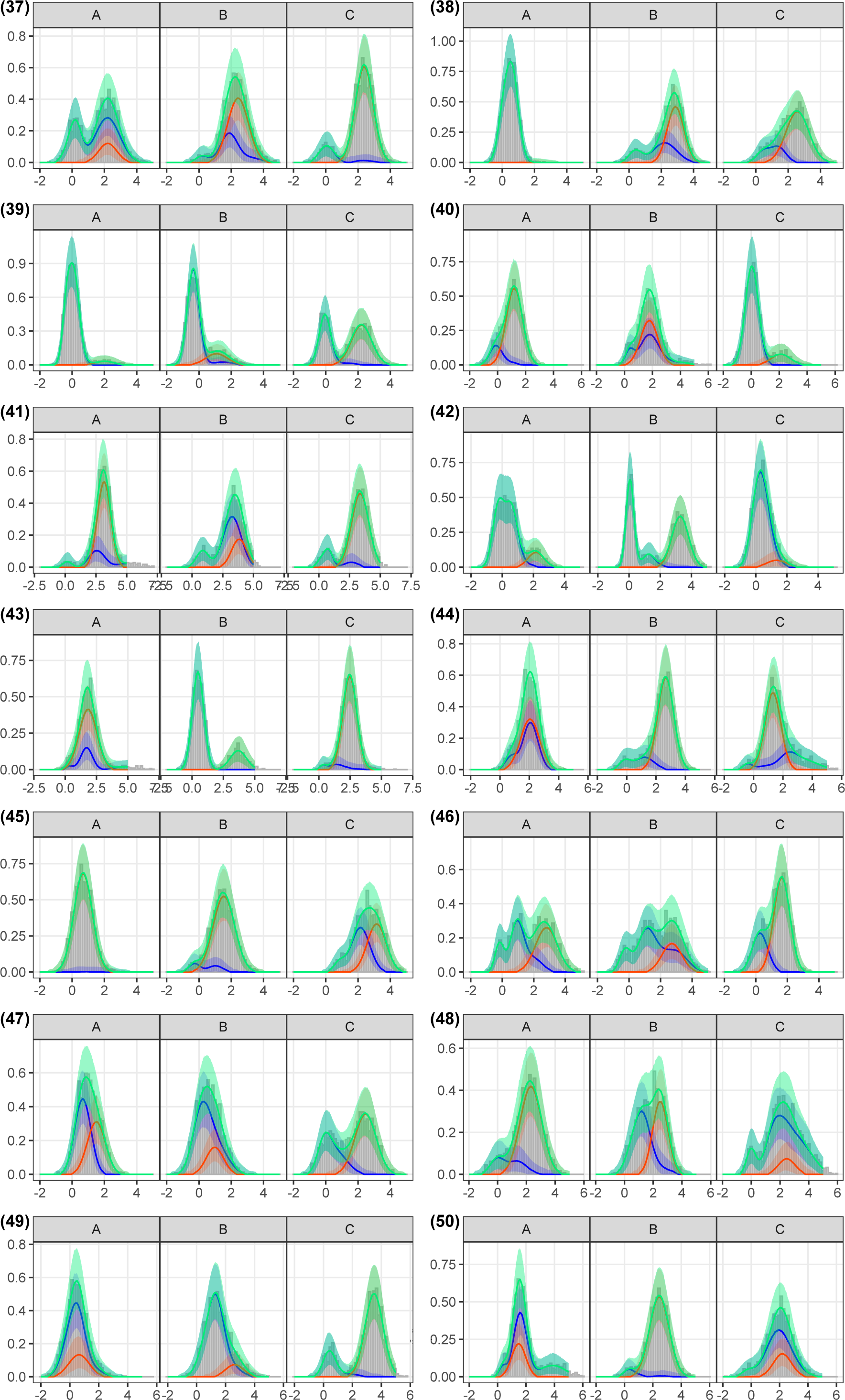
Model fits to 3D simulations. Grey bars show the density distribution of simulated antibody titers for each of the 50 simulations, for pathogens A, B and C. The lines and shaded ribbons show the median and 95%CrI model reconstructed titer distributions. Blue and orange indicate the reconstructed distribution of negative and positive titers to each pathogen respectively, while green shows the overall reconstructed titer distribution (combined across all components).

##### 3.2.3 Varying infection prevalence

In this section we fit two model versions to simulated data for 2 related pathogens where infection prevalence varies across 3 locations of equal sample size. We compared the performance of a base model that estimates only a single overall prevalence per pathogen and that of a location model that estimates pathogen-specific prevalence for each location. We found good consistency between estimated and true overall infection prevalence for both the base and location models with adjusted *R*^2^ values of 0.91 and 0.99, respectively. True values of overall infection prevalence values were within the estimated 95%CrIs for 80% and 100% of the base and location model estimates respectively as shown in Figure 15. In addition, 100% of location-specific prevalence estimates by the location model accurately recovered the true values (Figure 15). For the cross-reactivity parameters, *ϕ*, 91% and 96% estimates were accurately estimated by the base and location models respectively, shown in Figure 16. 84% and 92% of *ρ*_00_ and 84% and 94% of *ρ*_11_ estimates were accurately estimated by the base and location models respectively (Figure 16).

For *µ*_0_ parameters, true values were recovered for 92% and 95% of estimates across the 50 simulations, while for *µ*_1_ parameters true values were recovered in 92% and 98% of estimates for the base and location models respectively. In addition, 86% and 92% of *σ*_0_ true parameter values, and 92% and 96% of *σ*_1_ true parameter values were recovered by the base and location models respectively, shown in Figure 17. The location model outperformed the base model in all 50 simulations, as measured by DIC, WAIC and ELPD LOO. The median difference in DIC (base - location model) was 734.70 (range: 90.30 to 1967.9) and 729.73 (range: 90.76 to 1965.44) for WAIC. For the ELPD LOO metric the median difference (base - location model) was −364.9 (range: −982.71 to −45.38).

**Figure 15.**
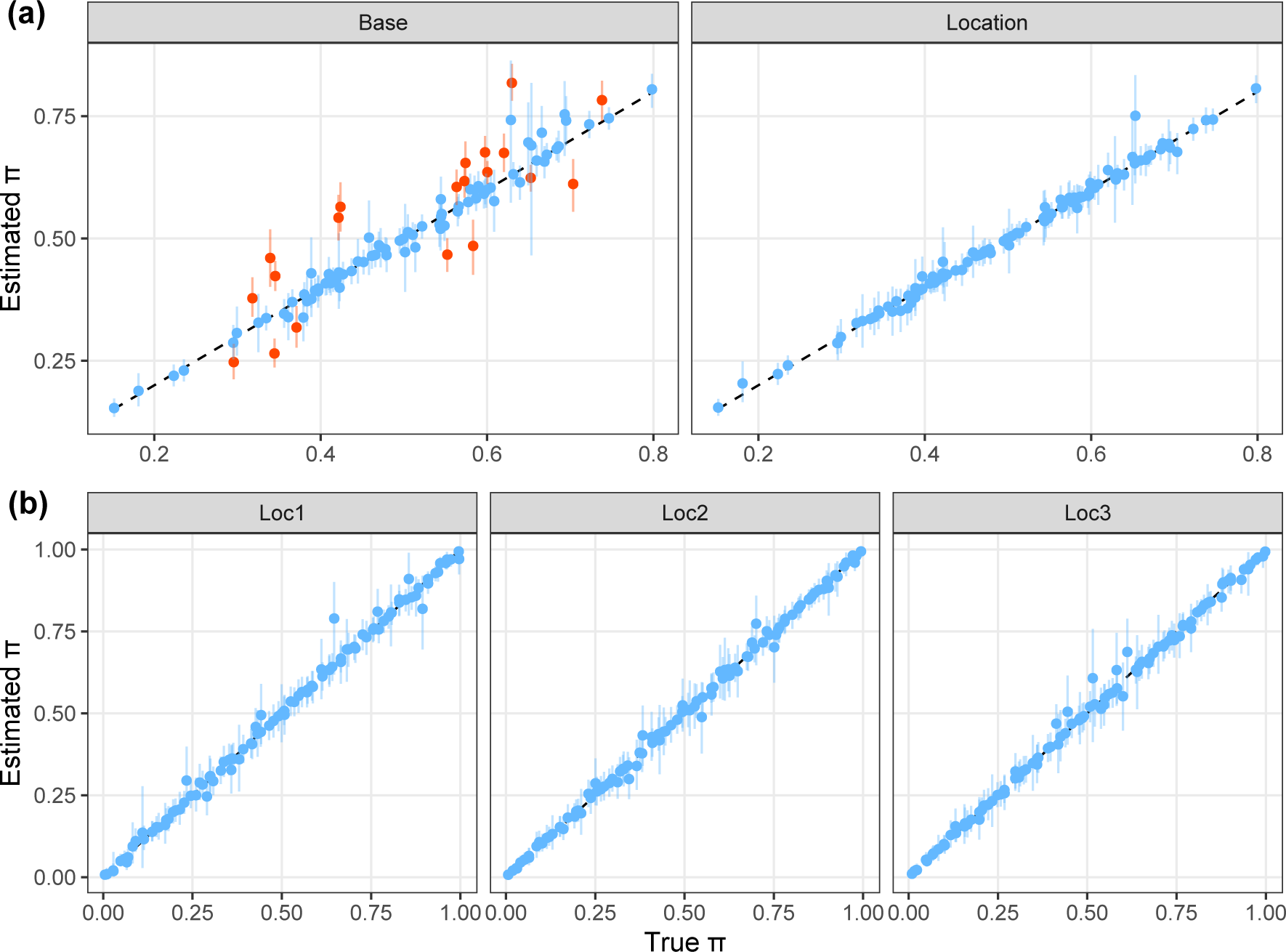
Recovery of location-specific infection prevalence estimates. Panel (a) shows the estimated vs true overall pathogen prevalence estimates from the base model and location model. Panel (b) shows the estimated vs true location-specific prevalence estimates from the location model. Coloured points and lines show the median and 95%CrI model estimates of the prevalence parameters, compared to the true values in 50 simulations. The black dashed line indicates where estimated values would equal the true values. Blue points and lines highlight the model estimates where the true value was contained within the 95%CrI and orange highlight those that did not contain the true value within estimated 95%CrI.

**Figure 16.**
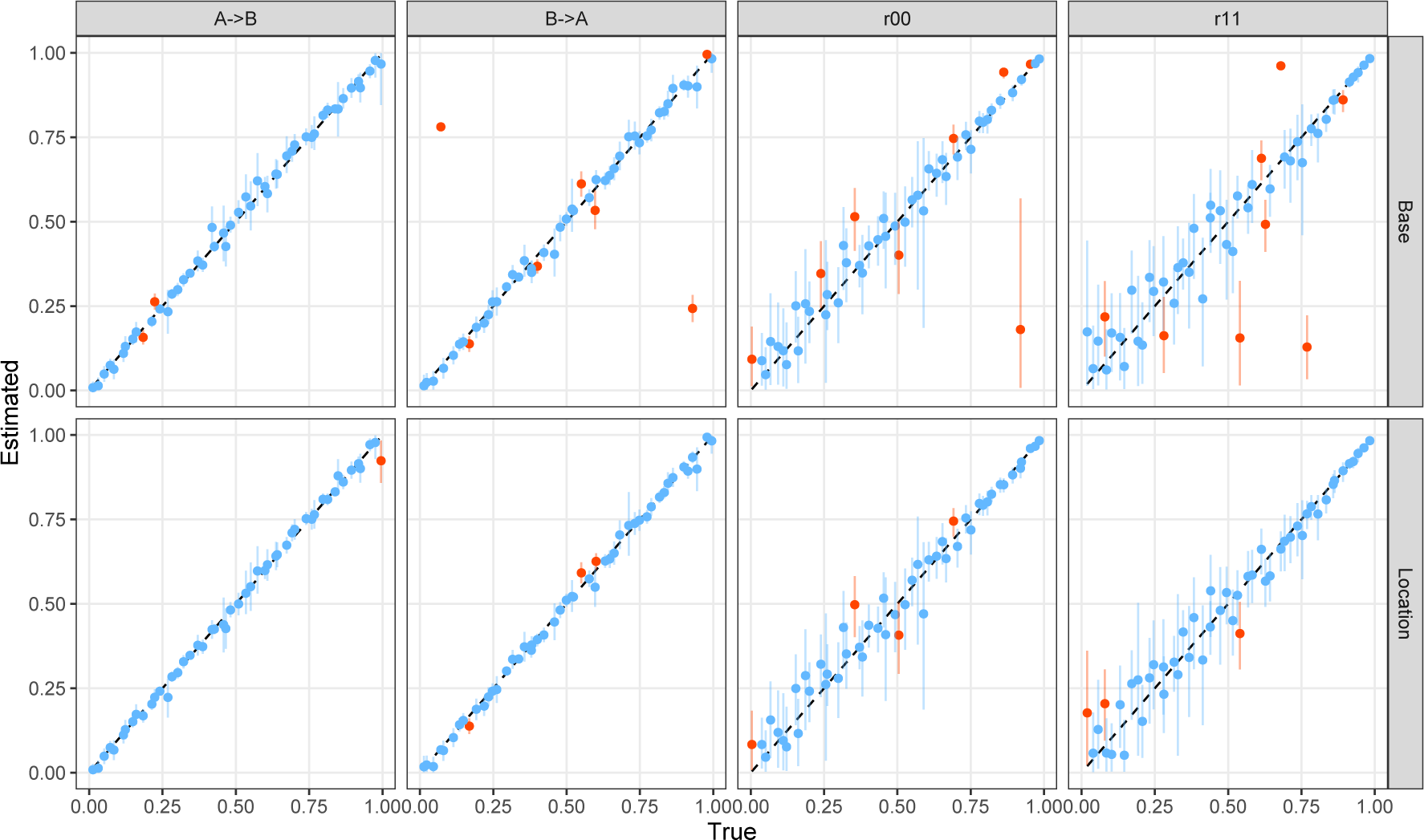
Recovery of cross-reactivity and correlation parameters with heterogeneous infection prevalence. Coloured points and lines show the median and 95%CrI model estimates of cross-reactivity and correlation parameters, compared to the true values in 50 simulations. The black dashed line indicates where estimated values would equal the true values. Blue points and lines highlight the model estimates where the true value was contained within the 95%CrI and orange highlight those that did not contain the true value within estimated 95%CrI. Panels A-¿B and B-¿A represent the estimates of relative titer increase from A to B and B to A, respectively. Panels r00 and r11 represent the correlation in the Gaussian distribution for infection statuses negative to both pathogens and positive to both pathogens, respectively. Row panels represent which model the estimates come from - the base model or location model.

**Figure 17.**
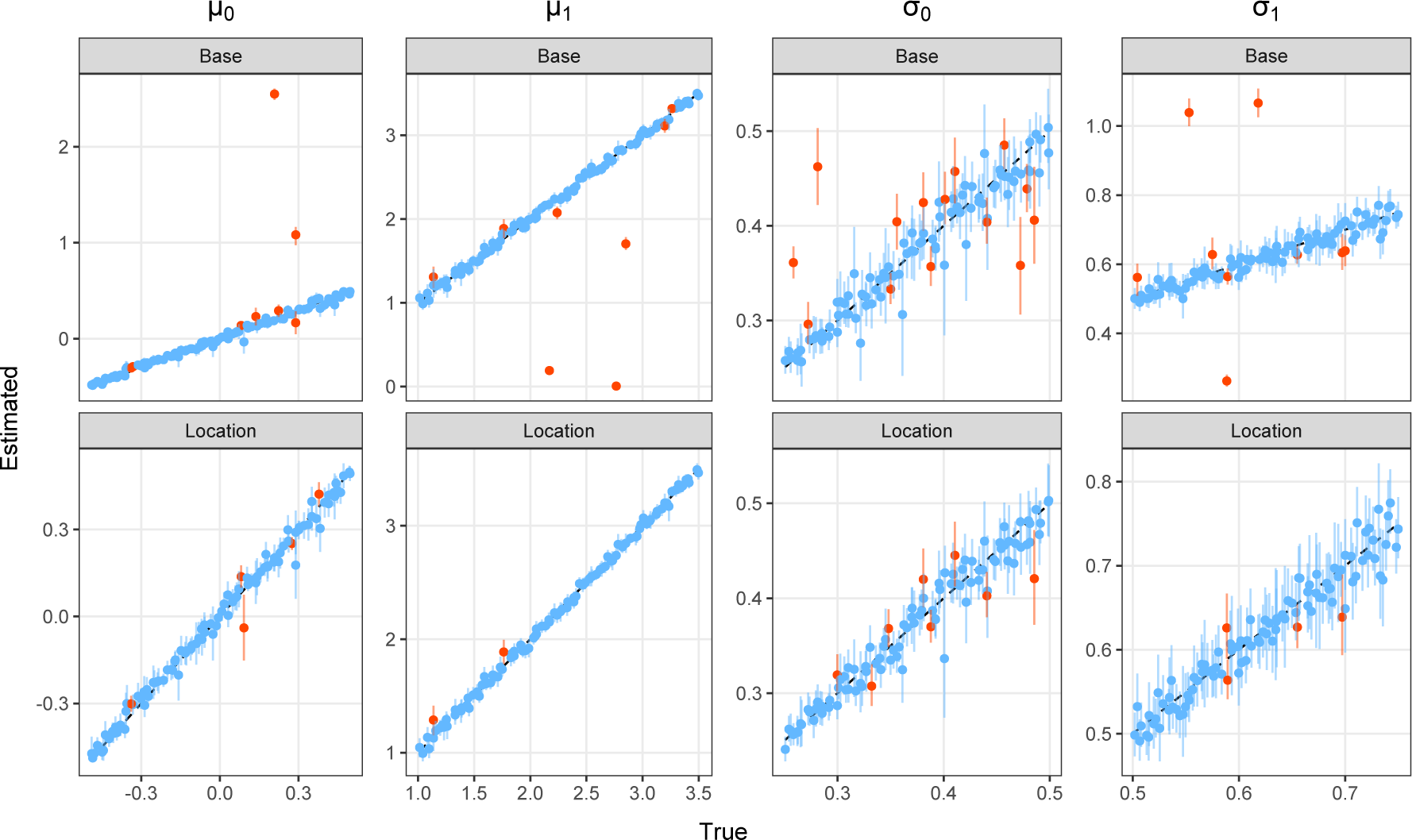
Estimated vs true Gaussian mean and standard deviation parameters from location and base models. Coloured points and lines show the median and 95%CrI model estimates of the *µ* and *σ* parameters, compared to the true values in 50 simulations. The black dashed line indicates where estimated values would equal the true values. Blue points and lines highlight the model estimates where the true value was contained within the 95%CrI and orange highlight those that did not contain the true value within estimated 95%CrI. Row panels represent which model the estimates come from - the base model or location model.

##### 3.2.4 1D model biases

To understand the levels of bias that can occur when cross-reactive antibodies are present but not accounted for, we fit classic single-dimension (1D) mixture models independently to each of the two pathogens from the same two pathogen system simulations of the previous section. Low consistency between estimated and true infection prevalence values, *π*, was observed from these models with an adjusted *R*^2^ of 0.40. True values of prevalence were contained within the 95%CrIs in 55% of estimates, shown in Figure 18, panel (a). We observed large overestimates of prevalence from the 1D models, coinciding with a general trend of underestimation of *µ*_1_ parameters and overestimation of *µ*_0_, shown in Figure 18, panel (b) and Figure 19. We found that the absolute bias in prevalence estimates is highest in simulations where both the level of prevalence of the unobserved pathogen is high and the level of cross-reactivity from infection with the unobserved pathogen against the observed pathogen is also high, shown in Figure 20. In contrast, high prevalence of the unobserved pathogen coupled with low cross-reactivity to the observed pathogen caused less bias in prevalence estimates. Similarly, high cross-reactivity from the unobserved to the observed pathogen coupled with low prevalence levels of the unobserved pathogen also resulted in reduced bias in prevalence estimates by the 1D model (Figure 20).

**Figure 18.**
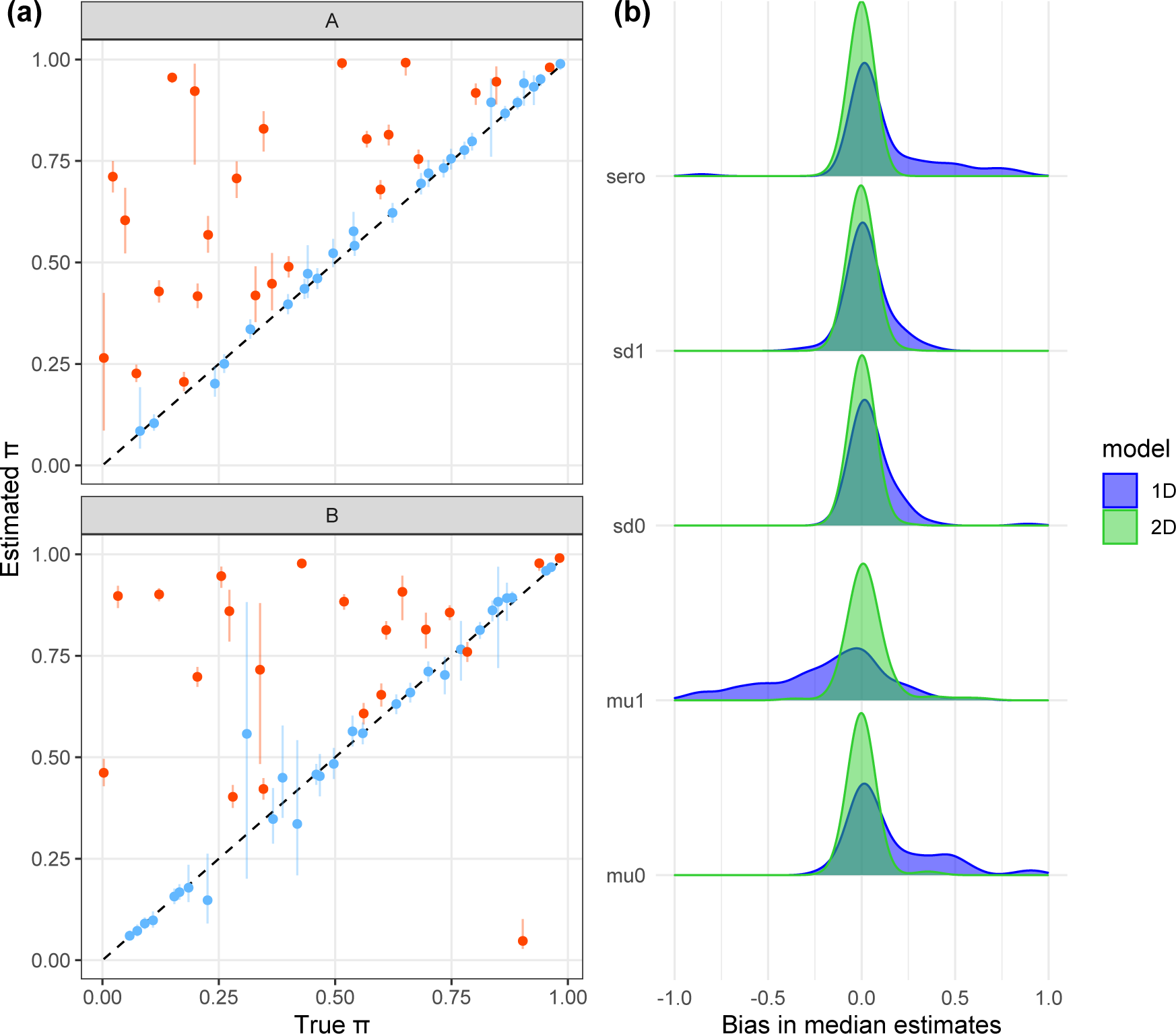
Bias in 1D model parameter estimates. Panel (a) shows the estimated vs true prevalence, *π*, from fitting classic 1D mixture models to 2D simulated data independently for pathogens A and B. Coloured points and lines show the median and 95%CrI model estimates of the *π*, compared to the true values in 50 simulations. The black dashed line indicates where estimated values would equal the true values. Blue points and lines highlight the model estimates where the true value was contained within the 95%CrI and orange highlight those that did not contain the true value within estimated 95%CrI. Panel (b) shows the density of bias (estimated - true) in model median estimates for all parameters for both the 1D (blue) and 2D models (green) for 50 simulations.

**Figure 19.**
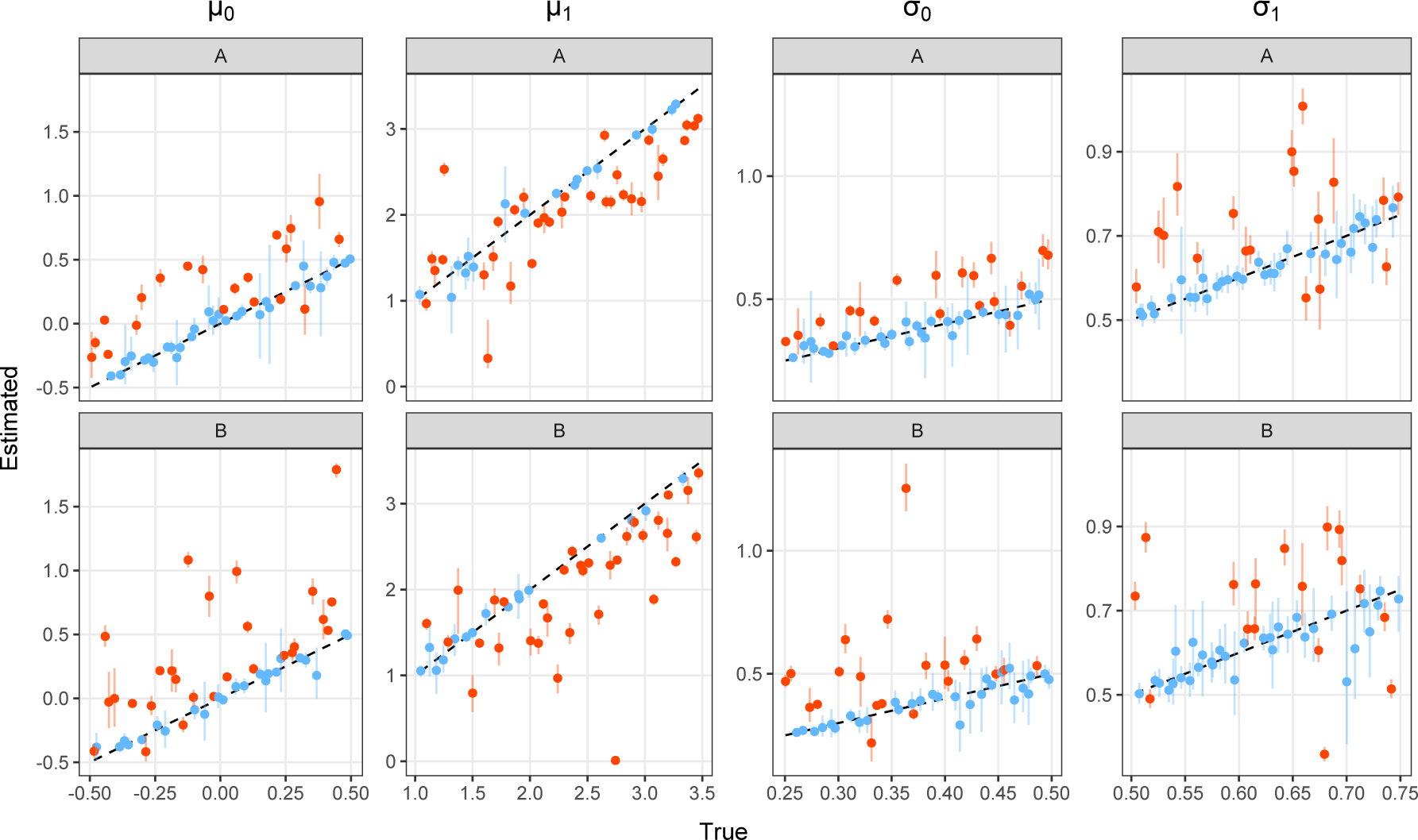
Estimated vs true Gaussian mean and standard deviation parameters from 1D models. Coloured points and lines show the median and 95%CrI model estimates of the *µ* and *σ* parameters, compared to the true values in 50 simulations. The black dashed line indicates where estimated values would equal the true values. Blue points and lines highlight the model estimates where the true value was contained within the 95%CrI and orange highlight those that did not contain the true value within estimated 95%CrI.

**Figure 20.**
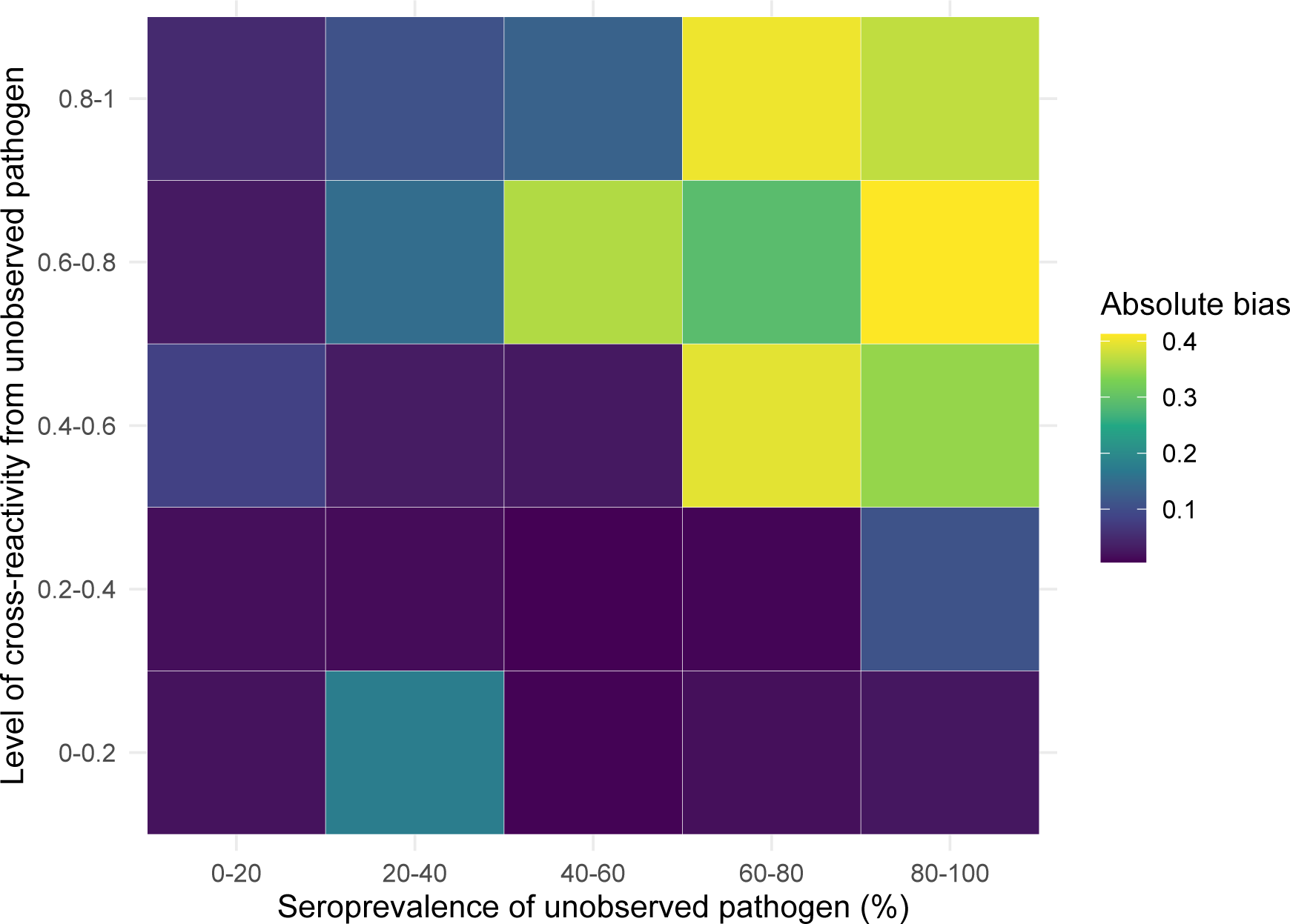
Bias in infection prevalence estimates from 1D models. Coloured tiles show the absolute bias in prevalence estimates by 1D models for varying levels of prevalence of the unobserved pathogen as well as varying cross-reactivity from infection with the unobserved pathogen against the observed pathogen.

##### 3.2.5 Determining pathogen presence vs absence

In this section we explored the ability of the model to differentiate between present and absent pathogens. Using the same 2 and 3 pathogen simulations as in the previous sections, we set the prevalence of one pathogen to zero and re-simulate the data with N=1,500. We then fit the same base model as before to each dataset as well as the “absent model” which assumes the absence of the truly absent pathogen. Only one 3D simulation was excluded from summary results due to non-convergence of model chains. High consistency between median estimates and true prevalence of the present pathogens was observed for both models with adjusted *R*^2^ values of 1.00 and 1.00 for the base and absent models respectively in both 2D and 3D simulations. Estimates of prevalence contained the true value within 95%CrIs for 100% of simulations for both the base and absent model in both 2D and 3D simulations, shown in Figure 21. For the absent pathogen, the base model estimates near zero infection prevalence for the majority of simulations, with a median estimate of 0.001 (range: *<*0.01-0.09) across simulations for the 2D system and a median estimate of 0.002 (range: *<*0.01-0.02) across the 3D simulations (Figure 21).

Cross reactivity parameters from the present pathogens to the absent pathogen were well recovered in both 2D and 3D simulations, shown in Figure 22. Cross reactivity estimates by the base model from the absent pathogens to the present pathogens, returned the prior in the majority of simulations (Figure 22). Correlation of negative titers, *ρ*_00_ was also well recovered in most simulations. Correlation of positive titers, *ρ*_11_ could only be well recovered in 3D simulations where a signal from the 2 present pathogens allowed it to be identified (Figure 22). In 2D simulations, as expected, the base model could not identify this parameter and returned the prior. High recovery of true Gaussian mean and standard deviation parameter values by the base and absent models were also observed for both 2D and 3D simulations. As expected, the base model returned the priors for *µ*_1_ and *σ*_1_ parameters that related to the absent pathogens.

To compare model performance between the base and absent models we calculated the difference in metrics between models (base model metric - absent model metric) as well as the LIPpp and LIPpc. The absent model was preferred to the base model by DIC in 45/50 2D simulations and 42/49 3D simulations, shown in Figure 23. Comparing model performance by WAIC, 44/50 simulations preferred the absent model in 2D simulations and 46/49 in 3D simulations. By ELPD LOO, 44/50 and 46/49 simulations preferred the absent model to the base. The median difference in log-likelihood values for 2D simulations was −0.74 (range: −1.19 - 5.23) and −0.79 (range: −1.58 − 5.21) in 3D simulations (Figure 23). Differences in DIC ranged from −2.33 to 8.01 with a median of 3.00 in 2D simulations, compared to a median of 3.43 in 3D simulations, ranging from −4.64 to 11.08. Median differences in WAIC were 1.72 (range: −6.93 - 3.05) and 1.85 (range: −6.43 - 4.93) in 2D and 3D simulations respectively. The median LIPpp for base compared to absent models across 2D simulations was 0.01 (range: −0.27 - 0.14) and 0.00 (range: −0.02 - 0.05) in 3D simulations, shown in Figure 23, panel B. The median LIPpc for base compared to absent models was −0.02 (range: −0.54 - 0.28) in 2D simulations and 0.00 (range: −0.02 - 0.06) in 3D simulations.

**Figure 21.**
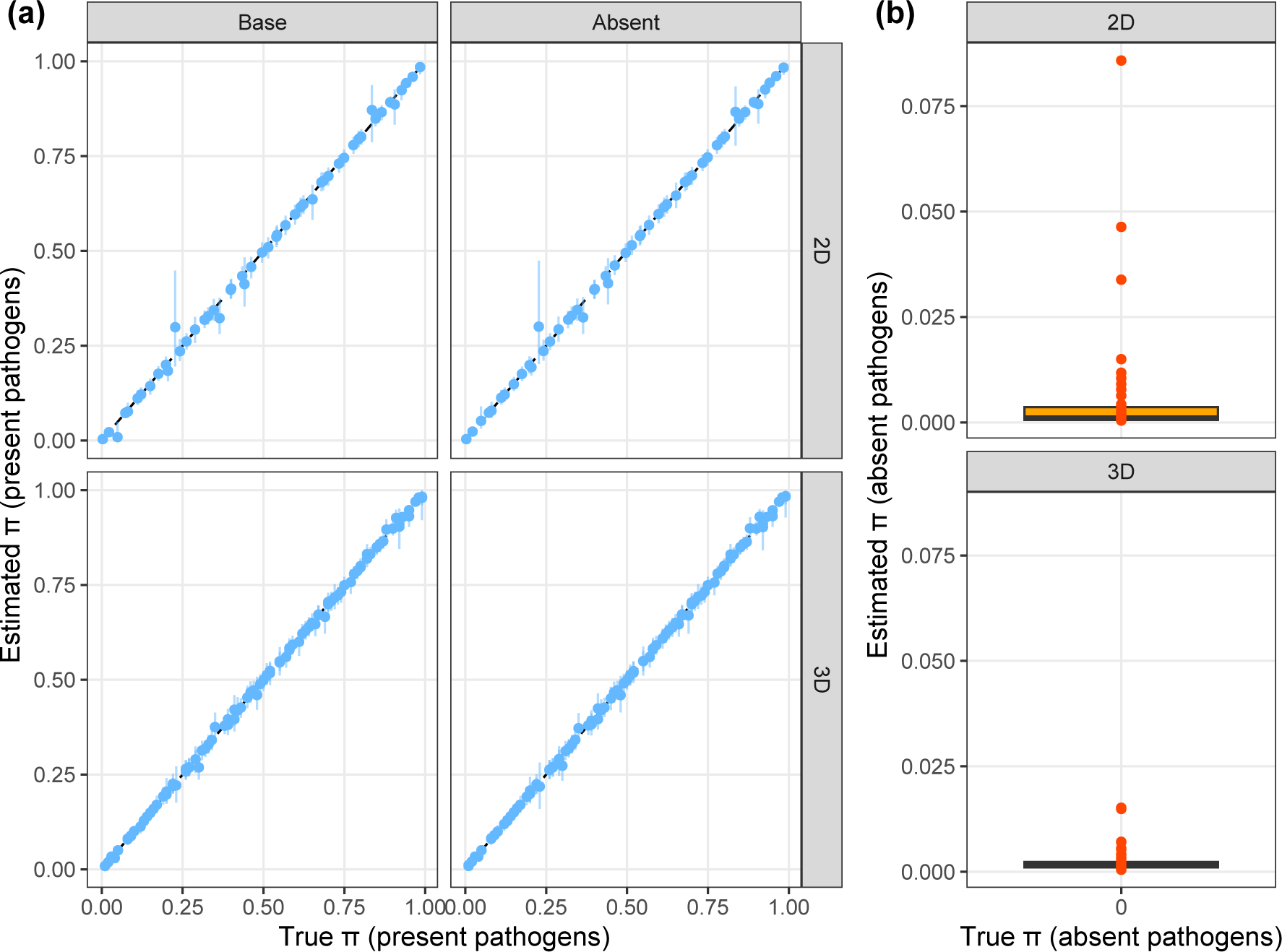
Recovery of infection prevalence parameters with absent pathogen. Panel (a) shows the estimated and true infection prevalence values *π* of present pathogens estimated by the base or absent model (column panels) for 2D and 3D simulations (row panels). Points and lines indicate median and 95%CrI estimates. Blue points and lines highlight the model estimates where the true value was contained within the 95%CrI and orange highlight those that did not contain the true value within estimated 95%CrI. The black dashed line indicates where estimated values would equal the true values. Panel (b) shows a box plot of median prevalence estimates by the base model for the absent pathogen (where true prevalence equals zero). Orange points represent the median estimates from individual simulations.

**Figure 22.**
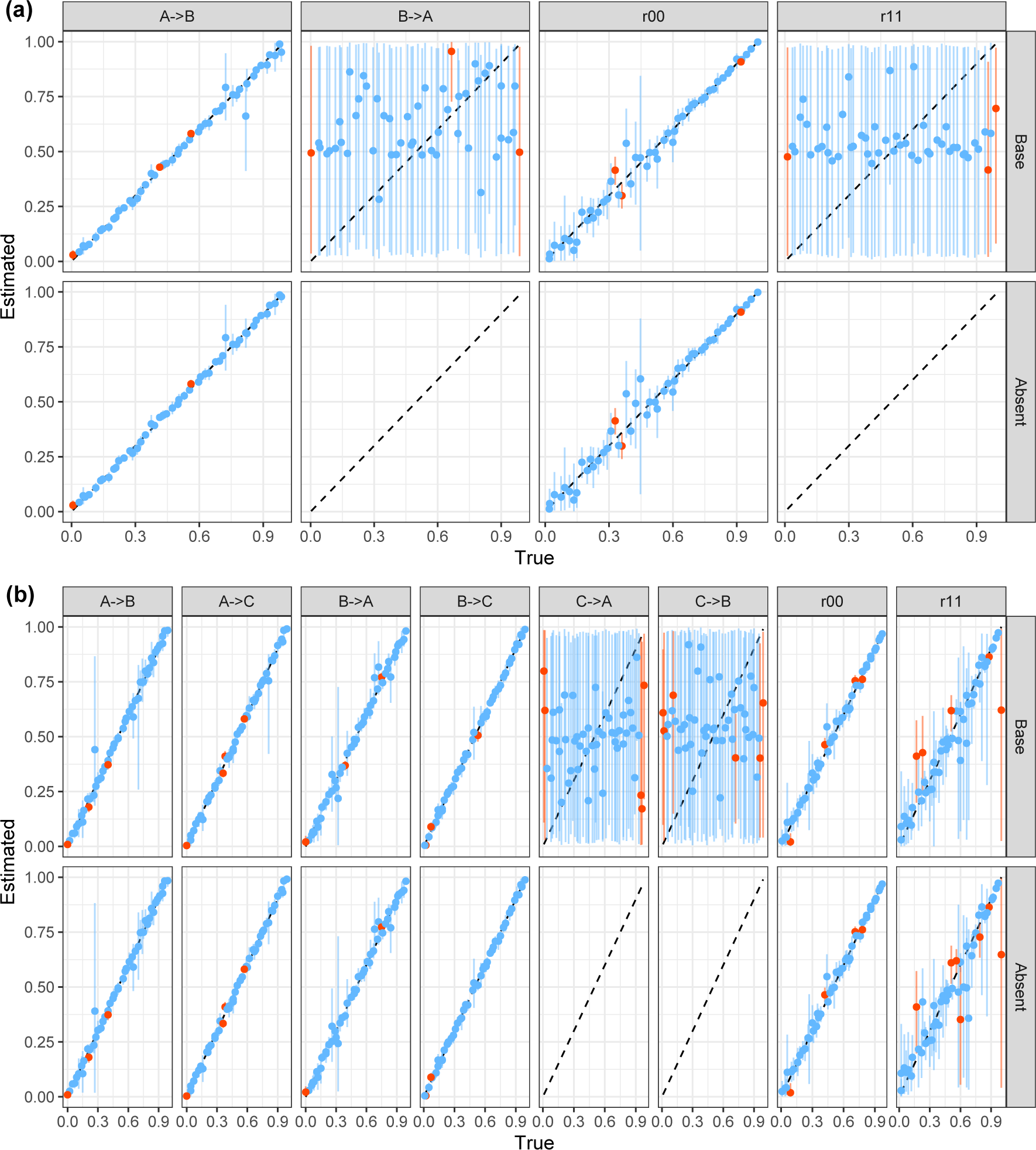
Recovery of cross reactivity and correlation parameters with absent pathogen. Panel (a) shows the estimated vs true values of cross reactivity parameters, *ϕ*, and correlation parameters, *ρ*, in 2D simulations. Panel (b) shows the same but for 3D simulations. Points and lines indicate median and 95%CrI estimates. Blue points and lines highlight the model estimates where the true value was contained within the 95%CrI and orange highlight those that did not contain the true value within estimated 95%CrI. The black dashed line indicates where estimated values would equal the true values. Column panels indicate individual parameters and row panels indicate the model used for fitting.

**Figure 23.**
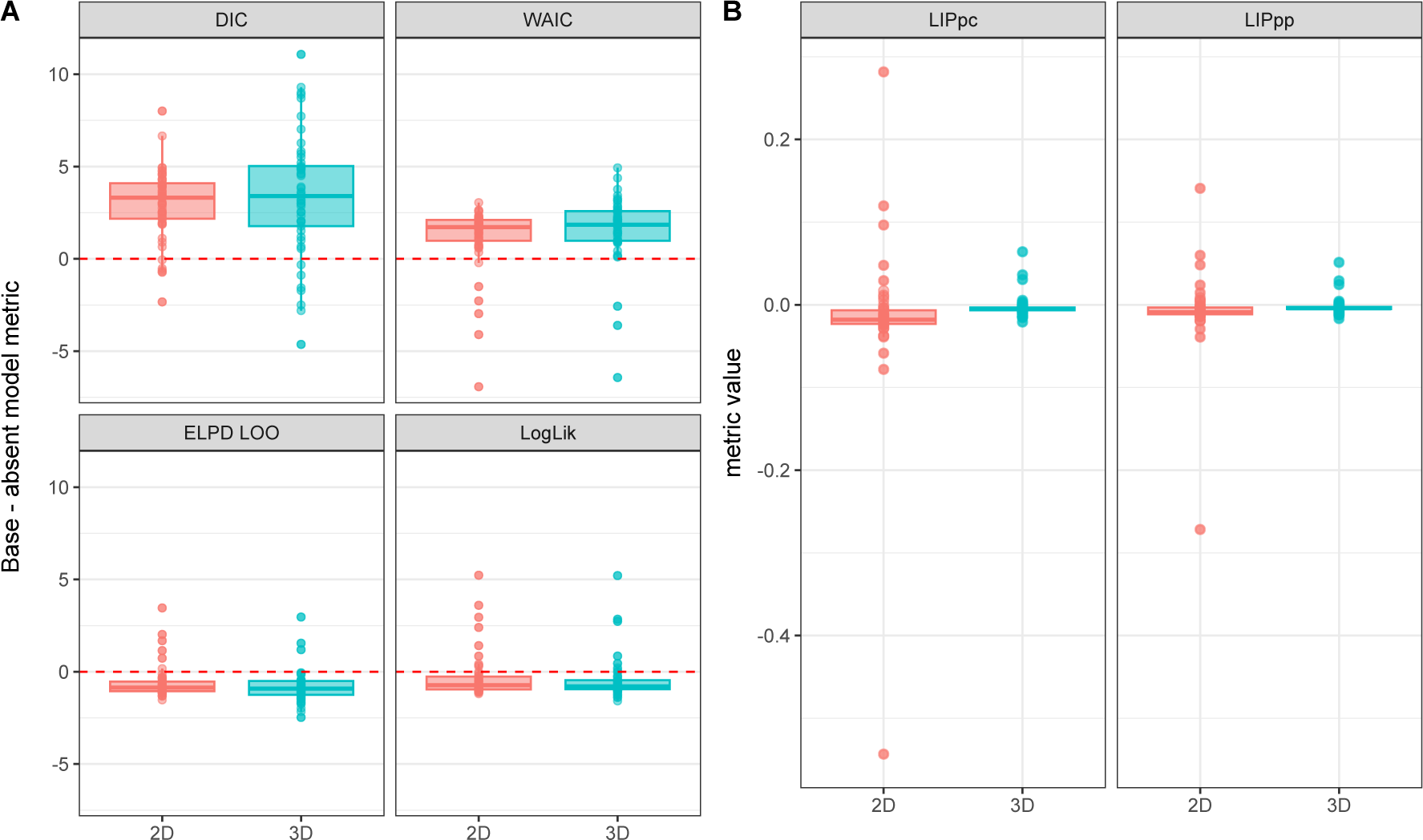
Absent vs Base model performance metrics. Panel A shows box plot summaries of the difference in model metrics (base minus absent model) for 2D and 3D simulations. Coloured boxes show the median and interquartile range of values while points show the individual values. Red dashed lines highlight a difference of zero, where base and absent model performances are equal. Panel B shows the likelihood increment percentage per parameter (LIPpp) and likelihood increment percentage per component (LIPpc) in 2D and 3D simulations. This calculates the percent increase in log likelihood per parameter or component for the base model compared to the simpler absent model.

